# Excess of de novo substitutions in the genomes of individuals with newly acquired germline *TP53* mutations

**DOI:** 10.64898/2026.11.19.26364643

**Authors:** Mathieu Parmentier, Matthew D. Montierth, Ruonan Li, Carlos Vera Recio, Gilda P. Chau, Bingshan Li, Rui Chen, Guillermina Lozano, Maxime Tarabichi, Wenyi Wang

## Abstract

Inherited defects in the tumour suppressor gene *TP53* cause Li-Fraumeni syndrome, a highly penetrant predisposition to childhood and adult cancers. About half of affected individuals carry a variant absent from both parents, and these are the patients missed by family-history-based testing criteria. Where and when such variants arise, in which parental gamete or after fertilisation, remains unresolved, as primary family data are scarce. We addressed this using whole-genome and linked-read sequencing of five family quartets, each comprising a proband with a confirmed newly arisen pathogenic *TP53* variant, a closely age-matched unaffected sibling, and both parents, drawn from a cohort of 429 families meeting clinical criteria for the syndrome; matching for age controls for the strong dependence of mutation counts on parental age. Every proband carried a distinct heterozygous missense change in the DNA-binding domain of p53, and read-based phasing traced four of five to the paternal chromosome. Genome-wide, probands carried a significant excess of newly arisen single-nucleotide changes relative to their siblings, which persisted after correction for parental age and was detected on the paternal chromosome. No comparable excess was found for insertions, deletions, structural rearrangements or repeat expansions, and the excess mutations matched the two clock-like processes operating in all cells rather than any distinct mechanism. These features are hard to reconcile with an event confined to the parental germ cells, and instead point to *TP53* variants that arise shortly after fertilisation and sweep the embryo, carrying neighbouring mutations with them. Consistent with such sweeps occurring throughout life, clonal *TP53* mutations in two large public tumour-genome collections ranked above other clonal driver mutations in cancer cell fraction within tumours, as expected when a cancer emerges within a pre-existing mutant patch of normal tissue. We propose that constitutional *TP53* variants and these mutant patches form one continuum, distinguished principally by timing: a sweep during the first cleavage divisions produces a variant present throughout the body, whereas the same advantage acting later confines the clone to one tissue. This would reframe a subset of Li-Fraumeni syndrome as an early developmental rather than strictly germline event, with implications for testing, counselling and surveillance.

## Introduction

*TP53*, encoding p53, plays a central role in cancer as a tumour suppressor gene^1^ and is the most frequently mutated gene in human malignancies. Germline pathogenic variants in *TP53* cause Li-Fraumeni syndrome (LFS), a highly penetrant autosomal-dominant cancer predisposition with lifetime cancer risk exceeding 75% in male and 90% in female carriers^2,3^. Population-scale exome analyses (gnomAD, n ≈ 138,632) place the carrier frequency of pathogenic germline *TP53* variants at ∼1 in 4,500. A substantial fraction of these pathogenic variants identified in patients with LFS are not inherited but arise *de novo*. Historical case series placed the *de novo* proportion at 7-20% of carriers^4^, but more recent series drawn from less heavily ascertained populations have revised this estimate sharply upward. For example, among 240 pediatric patients with osteosarcoma unselected for family history, 13 (5.4%) harbored a pathogenic or likely-pathogenic germline *TP53* variant, of which 6 out of 13 (46%) were confirmed *de novo* by parental sequencing - and *TP53* was the only gene with a recurrent *de novo* pathogenic variant in case-parent trios^5^. Another study applied a pedigree-based Mendelian model to 324 LFS families from four US cohorts estimated an approximately 1:1 ratio of *de novo* to inherited *TP53* mutations, with no major clinical-feature differences between *de novo* and inherited-mutation carriers^6^. Despite their clinical importance, the molecular origins and parental sources of *de novo TP53* mutations remain poorly characterized. Patients carrying them constitute an increasing share of opportunistically identified carriers, as routine tumour next-generation sequencing is coupled with germline interpretation^7^. Existing data on these families are thin. The largest single trio whole-exome study to confirm *de novo TP53* events identified six *de novo* pathogenic *TP53* carriers out of 95 trios^5^. Other trio-WES reports add *de novo TP53* mutations one family at a time^8–10^, accumulating to fewer than 25 in total. Trio whole-genome data are even scarcer^11,12^ with no published trio-WGS series of confirmed *de novo TP53* carriers. The complementary animal-model literature is similarly silent on origin: every published mouse model of germline *Trp53* loss or hotspot substitution^13–19^ uses an *inherited* germline allele in subsequent generations. No mouse model has been deliberately designed to recapitulate the *de novo* origin of germline *TP53* mutation, leaving open questions about parental gamete dynamics, paternal-age modifiers, and the origin and timing of *TP53* mutations that cannot be addressed in current systems.

Such *de novo* mutations are a general feature of human reproduction. Each child inherits, on average, ∼70 single-nucleotide variants not present in either parent, with roughly four out of five originating on the paternal allele^20,21^. The number observed in any individual offspring varies substantially around this mean: per-genome counts typically span ∼40-100 SNVs, with values exceeding 100 in children of older fathers and corresponding genome-wide rates of approximately 1.0-1.8 × 10 per nucleotide per generation^21–23^. The per-family rate at which DNMs accumulate with paternal age is itself heterogeneous, ranging from ∼0.2 to >3 additional mutations per year across multi-generation pedigrees, indicating substantial inter-family variability in germline mutational dynamics^24^. This paternal bias reflects the continuing divisions of spermatogonial stem cells throughout male reproductive life, which add ∼1-2 mutations per year of paternal age, in contrast to the largely arrested oocyte lineage of the female germline^24,25^. Studying these mutations is further complicated by a subset of apparent *de novo* mutations being not strictly *de novo* in the proband but arising from low-frequency parental gonadal or somatic mosaicism; deep sequencing of parental blood and sperm has identified mosaic origins in approximately 5-10% of *de novo* variants^26,27^. Family designs beyond the parent-child trio - quartets and three-or four-generation pedigrees - have become the principal tool for distinguishing true *de novo* events from variants inherited from a mosaic parent. The original family quartet of Roach et al. (2010)^28^ yielded one of the first direct human intergenerational mutation-rate estimates, and the design has since been scaled to the ∼2,700 quartets of the Simons Simplex Collection and follow-on WGS of 85 and 1,902 ASD quartets, in which the unaffected sibling provides a within-family control for *de novo* burden and artifact^29–31^. Therefore, we propose to apply the necessary sequencing design to study the origin of *de novo* pathogenic *TP53* mutations. In this study, we report whole-genome short- and linked-read sequencing of 5 age-matched family quartets in which the proband carries a confirmed *de novo* pathogenic *TP53* variant, with the goal of clarifying the origin of these variants.

## Results

### A unique cohort of 5 quartets controls for confounders and allows direct comparison of *de novo* mutation rates

Our initial dataset included 429 families, recruited at the University of Texas MD Anderson Cancer Center (MDACC) that meet the clinical LFS criteria (**Methods,**^32^). Among these, 138 families carried at least one *TP53* mutation whereas the remaining 291 families harboured no mutation and constituted the negative control cohort (**Figure 1A**). Based on the blood sample availability, we identified 28 family quartets, with each quartet consisting of both parents, proband and 1 sibling. These quartets were subsequently filtered according to the following criteria: a confirmed de novo *TP53* mutation in the proband; a 4-year maximum age difference between the proband and the sibling; the paternal ages at the child’s birth are within 10 years; and no record of any cancer in parents or siblings. We used these criteria to ensure the comparability of de novo mutation rates between and across the proband-sibling pairs. A total of 6 family quartets satisfied all criteria (**Figure 1A**). One family was then further excluded (m1) due to the presence of an additional germline pathogenic mutation in the proband (**Supplementary Figure S1**), leading to a cohort of 5 quartets (**Figure 1B**) with age-matched probands and siblings, and a similar range of parental ages at child birth (**Figure 1C**). DNM rates scale linearly with parental age, with a faster increase for paternal age^20,33^. In our cohort, the average age difference between proband and sibling was 2.2 years ([-2, +4] years; **Figure 1C**) with no systematic bias, limiting the potential impact of parental age differences on DNM rates. Of note, each patient developed at least 2 cancers characteristic of LFS, including osteosarcoma, breast cancers and other sarcomas (**Figure 1D-E**). We obtained whole-genome sequencing (WGS) data (35.7X median coverage depth) and linked-read sequencing (LRS) data (7.35X median coverage depth) for 6 family quartets, that is 1 proband with de novo *TP53* mutations detected in the blood; 1 age-matched sibling; their mother and father; and 2 trios as controls (1 proband with inherited TP53 mutations; their mother and father) (**Table 1 - 2, Supplementary Figure S2**).

**Figure 1:**
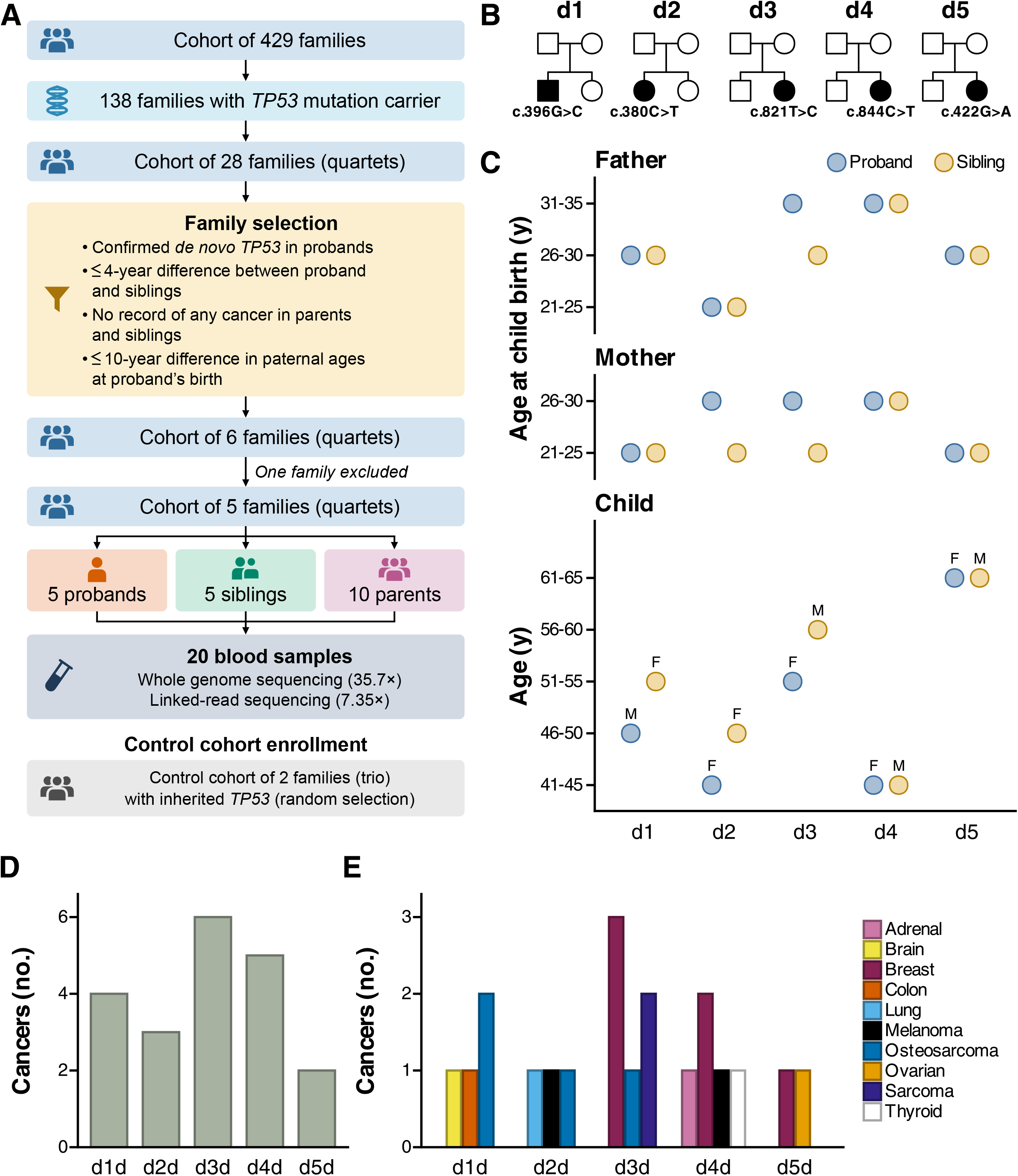
Study design of Li-Fraumeni cohort. A) Strategy of family selection. Families were selected according to the corresponding criteria: - Family of quartet (proband, sibling and parents), - Confirmed de novo TP53 mutation in probands, - Germline DNA samples for each quartet, - Less than 4-year difference between proband and sibling, - No record of any cancer in parents and siblings, - Maximum 10-year difference in paternal ages at proband’s birth. Family m1 was excluded due to a deletion variant in the PALB2 gene in the proband. Blood samples were processed using Whole genome sequencing (WGS) and Linked-read sequencing (LRS). B) Family pedigree. Probands are indicated by an arrow. C) Comparison of paternal, maternal age at birth, proband and sibling’s age in each family. The average age difference between proband and sibling is 2.2 years. D) Repartition of cancer types across Li-Fraumeni patients.

**Table 1:** General description of the cohort of individuals. Each person was sequenced using short-read whole-genome sequencing (WGS), with their familial status, date of birth, TP53 mutation type, and sequencing depth reported according to quantiles.

| Family | Family Label | Symbol | Member | TP53 | Read depth<br>(0.9 quantile) | Read depth<br>(0.5 quantile) | Read depth<br>(0.1 quantile) |
|---|---|---|---|---|---|---|---|
| Familial 1 | F1 | f1f | Father | / | 49 | 40 | 30 |
| Familial 1 | F1 | f1m | Mother | / | 48 | 39 | 30 |
| Familial 1 | F1 | f1d | Proband | hereditary | 52 | 42 | 32 |
| Familial 2 | F2 | f2f | Father | / | 49 | 40 | 30 |
| Familial 2 | F2 | f2m | Mother | / | 47 | 38 | 30 |
| Familial 2 | F2 | f2d | Proband | hereditary | 50 | 40 | 32 |
| De novo 5 | D5 | d5f | Father | / | 47 | 38 | 28 |
| De novo 5 | D5 | d5m | Mother | / | 46 | 37 | 29 |
| De novo 5 | D5 | d5d | Proband | de novo | 50 | 40 | 32 |
| De novo 5 | D5 | d5s | Sibling | / | 49 | 40 | 30 |
| De novo 4 | D4 | d4f | Father | / | 49 | 39 | 29 |
| De novo 4 | D4 | d4m | Mother | / | 48 | 38 | 29 |
| De novo 4 | D4 | d4d | Proband | de novo | 52 | 42 | 33 |
| De novo 4 | D4 | d4s | Sibling | / | 45 | 36 | 27 |
| De novo 3 | D3 | d3f | Father | / | 52 | 42 | 32 |
| De novo 3 | D3 | d3m | Mother | / | 48 | 39 | 30 |
| De novo 3 | D3 | d3d | Proband | de novo | 48 | 39 | 30 |
| De novo 3 | D3 | d3s | Sibling | / | 50 | 40 | 29 |
| De novo 2 | D2 | d2f | Father | / | 49 | 39 | 30 |
| De novo 2 | D2 | d2m | Mother | / | 48 | 39 | 30 |
| De novo 2 | D2 | d2d | Proband | de novo | 52 | 42 | 32 |
| De novo 2 | D2 | d2s | Sibling | / | 47 | 38 | 30 |
| De novo 1 | D1 | d1f | Father | / | 52 | 42 | 31 |
| De novo 1 | D1 | d1m | Mother | / | 50 | 40 | 31 |
| De novo 1 | D1 | d1d | Proband | de novo | 52 | 42 | 31 |
| De novo 1 | D1 | d1s | Sibling | / | 49 | 39 | 30 |
| Mosaic 1 | M1 | m1f | Father | / | 47 | 37 | 25 |
| Mosaic 1 | M1 | m1m | Mother | / | 49 | 40 | 30 |
| Mosaic 1 | M1 | m1d | Proband | de novo | 49 | 39 | 29 |
| Mosaic 1 | M1 | m1s | Sibling | / | 48 | 38 | 28 |

**Table 2:** Parental age at birth and current age of offspring (n = 14). All ages are grouped in 5-year intervals.

| child | Father age at birth - interval | Mother age at birth - interval | Child age - interval |
|---|---|---|---|
| d1d | 26-30 | 21-25 | 46-50 |
| d1s | 26-30 | 21-25 | 51-55 |
| d2d | 21-25 | 26-30 | 41-45 |
| d2s | 21-25 | 21-25 | 46-50 |
| d3d | 31-35 | 26-30 | 51-55 |
| d3s | 26-30 | 21-25 | 56-60 |
| d4d | 31-35 | 26-30 | 41-45 |
| d4s | 31-35 | 26-30 | 41-45 |
| d5d | 26-30 | 21-25 | 61-65 |
| d5s | 26-30 | 21-25 | 61-65 |
| f1d | 26-30 | 21-25 | 66-70 |
| f2d | 36-40 | 31-35 | 41-45 |
| m1d | 31-35 | 26-30 | 46-50 |
| m1s | 31-35 | 26-30 | 46-50 |

First, we called mutations genome-wide, genotyped all our samples for *TP53* mutations, and screened for potential pathogenic variants (**Methods**). This showed that each proband harboured a distinct missense mutation in exon 5 or exon 8 of *TP53*, which encodes the DNA-binding domain (**Figure 2A**). Among these, 4 out of the 5 missense mutations were classified as pathogenic, according to the American College of Medical Genetics and Genomics guidelines; whereas the mutation c.396G>C (p.Lys132Asn) was classified as Likely pathogenic. Of note, the proband from the d4 family harboured the missense mutation c.844C>T (p.Arg282Trp), one of the hotspot mutations in LFS. In each family, the *TP53* variant was exclusive to the proband and had an allele-frequency of ∼0.5 while no other pathogenic variants were detected. In the excluded family, the *TP53* variant had an allele frequency of 0.2 (10 out 49 reads; significantly below 0.5 - P_Binomial_ = 3.8e-05), suggesting a mosaic or subclonal *TP53* variant in the blood lineage. Additionally, the proband also carried a clonal heterozygous pathogenic deletion variant in the *PALB2* gene (18 out of 40 reads), known to be related to phenotype associated with higher cancer risks (**Supplementary Figure S1**), which altogether led to its exclusion from our analyses.

**Figure 2:**
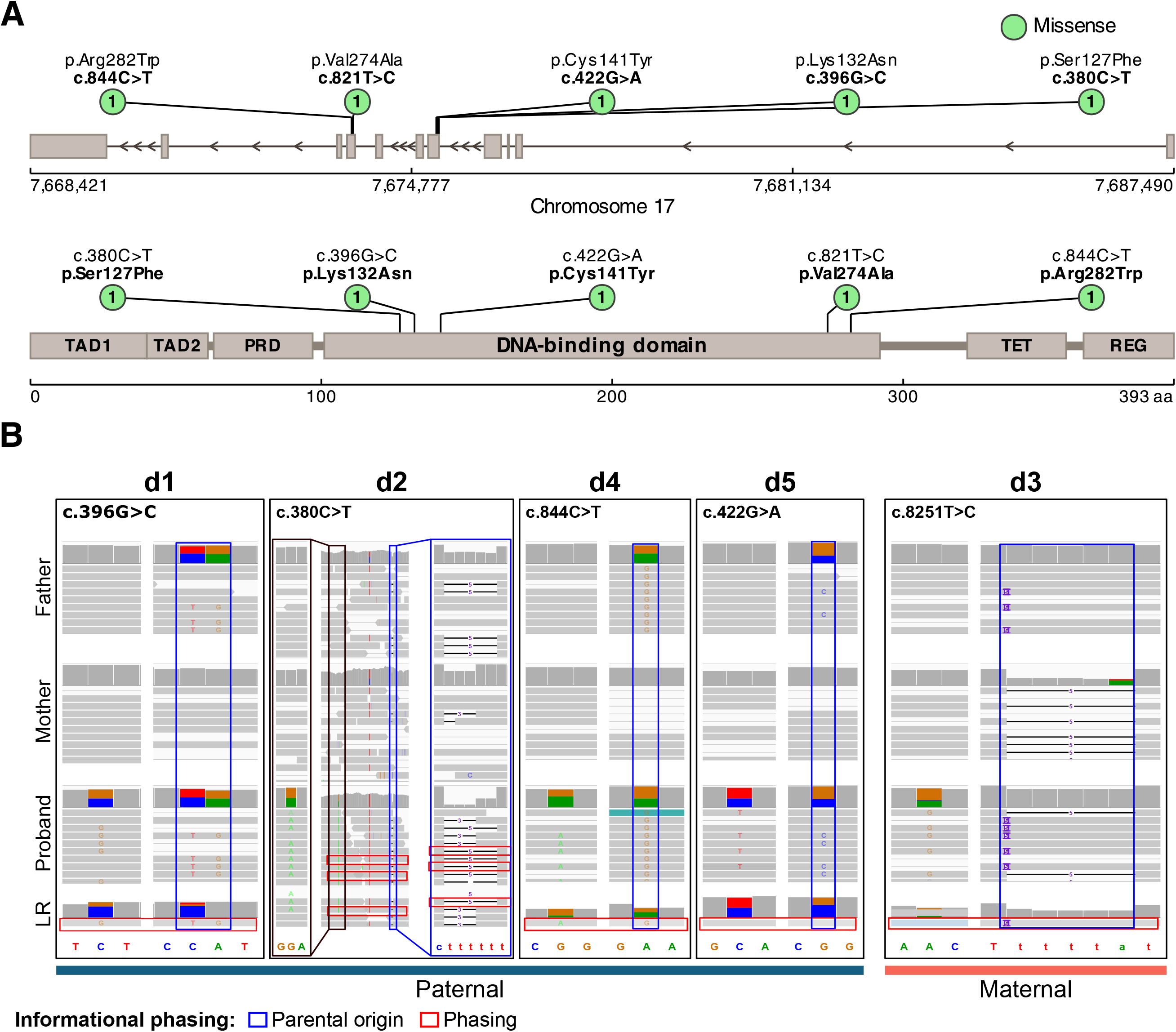
Identification of TP53 mutations in the Li-Fraumeni cohort. A) Lollipop plots displaying mutations in TP53 gene and protein across the cohort. Gray tick marks indicate TP53 mutations classified as ‘pathogenic’ or ‘likely pathogenic’ in association with Li-Fraumeni syndrome in ClinVar. Each patient harbors a missense mutation in exon 4 or 7 and within the DNA binding domain of TP53 protein. (B) Coverage tracks highlighting TP53 mutation phasing with both Whole-genome (WGS) and Linked-read sequencing (LRS) and with WGS only (C). TP53 mutation is displayed on the left of each coverage track. The TP53 mutation was phased in 4 families to a paternal variant and a maternal in d3 family.

To avoid any bias in our analysis due to potential additional pathogenic mutations, we screened our cohort for pathogenic germline mutations with a 3-out-of-3 consensus mutation calling pipeline (**Methods**). We detected a total of **7,786,358 unique** SNVs and **2,182,035** INDELs, with an average value of **3,374,984** and **727,023** per sample, respectively. None of these mutations were detected as pathogenic or likely pathogenic **(Supplementary Figure S3).**

Using both WGS and LRS data, we next phased *TP53* mutations with SNP-in-read phasing to nearby heterozygous SNPs to identify the parental copy carrying the mutation (**Methods**). Combination data from both short- and linked-reads, we could phase the *TP53* variant to a parent-specific genotype in all of the 5 quartet families: 4 out of 5 variants were phased to the paternal copy, while the variant in the d3 family was phased to the maternal copy (**Figure 2B**).

### A significant excess of de novo substitutions in *TP53*-mutated probands

To obtain high-confidence DNM calls, we implemented and ran a 2-out-of-4 consensus substitution and indel calling pipeline (**Methods**). This identified 735 de novo substitutions and 117 de novo indels in the cohort, with an average of 93 per proband and 77.4 per sibling (**Supplemental Figure S4**; **Table 3**). Among these, only *TP53* DNMs were classified as pathogenic and likely pathogenic (**Supplementary Figure S5**).

**Table 3:** Summary of de novo SNVs across the cohort, with their phaseability and parental origin.

| Family | D1 |  | D2 |  | D3 |  | D4 |  | D5 |  |
|---|---|---|---|---|---|---|---|---|---|---|
| Child | Prob. | Sibl. | Prob. | Sibl. | Prob. | Sibl. | Prob. | Sibl. | Prob. | Sibl. |
| de novo SNV * | **82** | **78** | **72** | **50** | **90** | **70** | **91** | **77** | **71** | **54** |
| Phaseable SNV | 37 | 43 | 28 | 24 | 40 | 38 | 42 | 42 | 37 | 25 |
| SNV with origin known | 32 | 33 | 23 | 20 | 35 | 33 | 30 | 33 | 31 | 23 |
| SNV on the paternal copy | 21 | 25 | 17 | 14 | 29 | 29 | 23 | 24 | 27 | 19 |
| Estimated SNV (Paternal copy) | 54 | 59 | 53 | 35 | 75 | 62 | 70 | 56 | 62 | 45 |
| SNV on the maternal copy | 11 | 8 | 6 | 6 | 6 | 4 | 7 | 9 | 4 | 4 |
| Estimated SNV (maternal copy) | 28 | 19 | 19 | 15 | 15 | 8 | 21 | 21 | 9 | 9 |

To validate these, we ran Sanger sequencing on **35 DNMs** for which we could design primers, on blood samples of parent-child trio, leading to a validation rate of **100% (**35 out of 35 validated DNMs; **Figure 3A)**.

**Figure 3:**
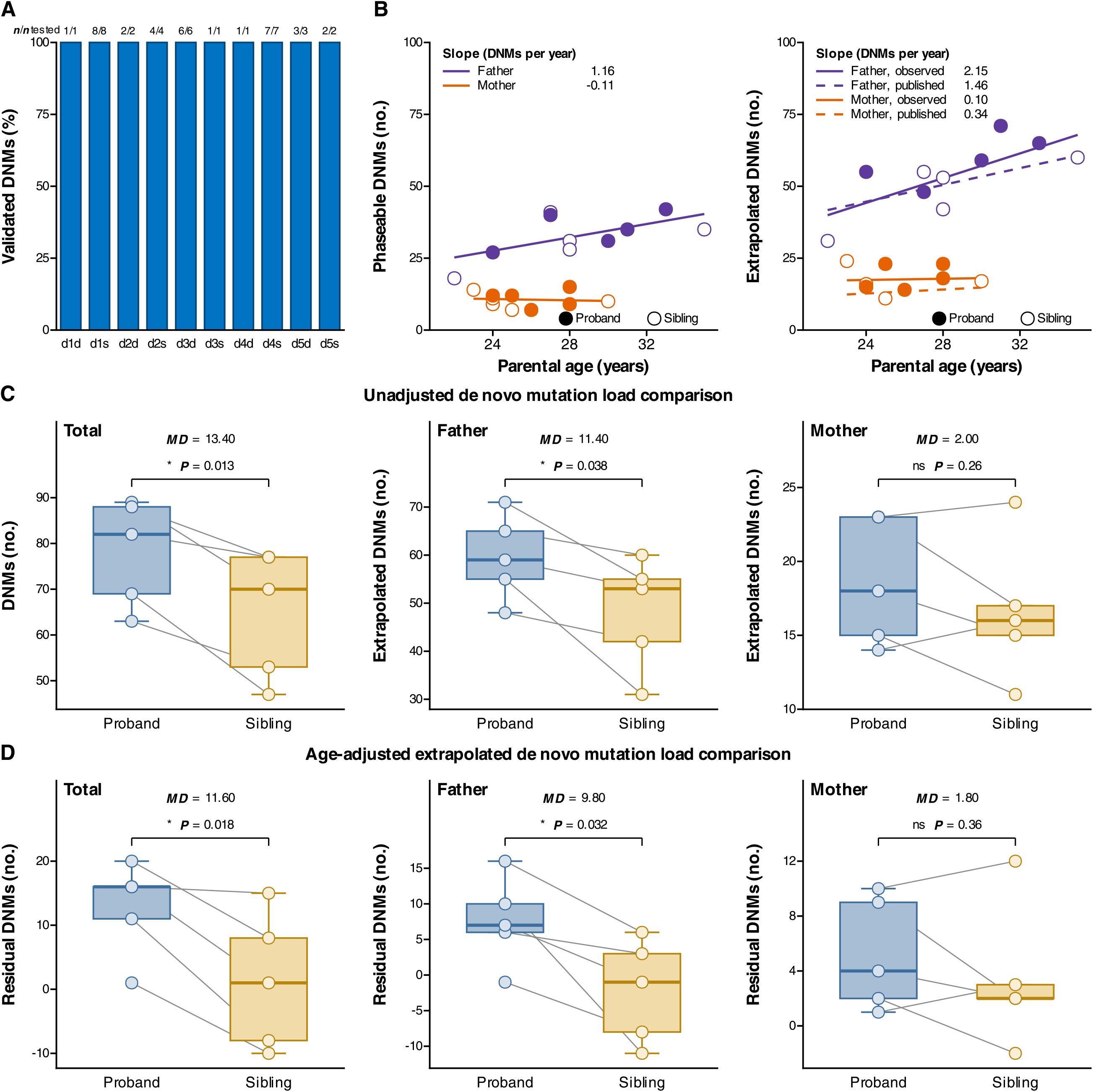
Excess of de novo substitutions in probands from both parental copies. A) DNM SNVs validated by sanger sequencing and detected in the consensus pipeline. B) Mutation rate inferred as a function of paternal and maternal age with phaseable DNM count (Paternal R² = 0.378, *P =* 0.059; Maternal: R² = 0.009, *P =* 0.796), and extrapolated DNM count (Paternal R² = 0.54, *P = 0.0154*; Maternal: R² = 0.003, P *=* 0.883) in autosomes. C) Comparison of total and extrapolated DNM count in autosomes between probands and siblings (Total: *P* = 0.013, MD = 13.40; Maternal: *P* = 0.258, MD = 2.00; Paternal: *P* = 0.038, MD = 11.40, paired t-test). D) Comparison of residuals DNM in autosomes between probands and siblings (Total: *P* = 0.018, MD = 11.60; Maternal: *P* = 0.36, MD = 1.80; Paternal: *P* = 0.032, MD = 9.80, paired t-test). Residuals were computed as DNMs_theoretical_ – DNMs_extrapolated_, MD = mean difference between probands and siblings. NS = not significant (P > 0.05). Each star symbol (*) indicates P < 0.05.

Our results revealed a significant excess of de-novo substitutions on autosomes in probands relative to their age-matched control siblings (MD = 13.40, paired t-test **P=0.009**; **Figure 3C**) but no detectable excess in indels (**Supplementary Figure S4 A**).

We reasoned that this excess could have occurred either pre-fertilisation, or post-fertilisation in the few first cell divisions. In the first case, there would be a parent-of-origin effect for the excess; whereas in the latter there would likely not.

To tackle this question, we phased mutations on autosomes to the nearby heterozygous polymorphic variants (SNPs and INDEL) to infer their parent of origin (**Methods**). Combining the high-depth short reads with the linked reads, we were able to determine the origin of a median of 58% of de novo mutations, with the proportion of variants with an assigned origin ranging from 49.2% to 82% across children. This demonstrated an average of 77% mutations of paternal origin and 22% mutations of maternal origin, in line with estimates from a large trio cohort from Iceland^21^ (**Figure 3B; Methods**). However, we could not detect an excess in the number of mutations from either parent between probands and siblings (**Supplementary Figure S4 B**; Paternal copy: *P*=0.25; Maternal copy: *P*=0.66), potentially due to the low number of phased mutations and lack of power. Because not all mutations are attributable to a parent of origin, we extrapolated the total number of mutations coming from each parent by multiplying the total mutation count by the proportion of origin-determined mutations attributed to each parent (**Figure 3C**). This makes the reasonable assumption that there would be no bias in parent of origin between phaseable and non-phaseable mutations. Here, we observed a significant excess in mutations assigned to the father allele (*P*=0.04) whereas no excess was detectable in mutations on the mother allele (*P*=0.26).

To compare how probands and siblings deviated from the expected DNM counts, we next computed the DNM residuals as the difference between the extrapolated mutation counts and the mutation counts expected given the age of the parents^21^ (**Methods**; **Figure 3D**).

We observed significantly higher total and paternal residuals in probands compared to siblings (paired *t*-test, Total; *P* = 0.018, Paternal: *P* = 0.032) and higher than the average expected counts^21^, but no difference for maternal residuals (*P* = 0.36).

Altogether, we find significantly different numbers of DNM between probands and siblings, even when accounting for the age of the parents, suggesting a higher load specifically in the proband carrying a deleterious DNM in *TP53*. The effect is also significant in the mutations specifically on the paternal allele, but not detectable in mutations falling on the maternal allele. Due to low power, from DNM counts alone, we cannot distinguish between a pre-fertilisation or post-fertilisation effect.

### Similar rates of indels, structural variants and short tandem repeats and known underlying mutational processes

Next, we tested whether a similar excess could be seen in structural variants (SV), or short tandem repeats (STR). First, we implemented a consensus de novo mutation calling pipeline for SV (**Methods**). We identified a single de novo SV in both the proband from family d2 and sibling from family d3 (**Figure 4A**). This is in line with the previously reported low detection rate of de novo SV by short-read WGS^34^.

**Figure 4:**
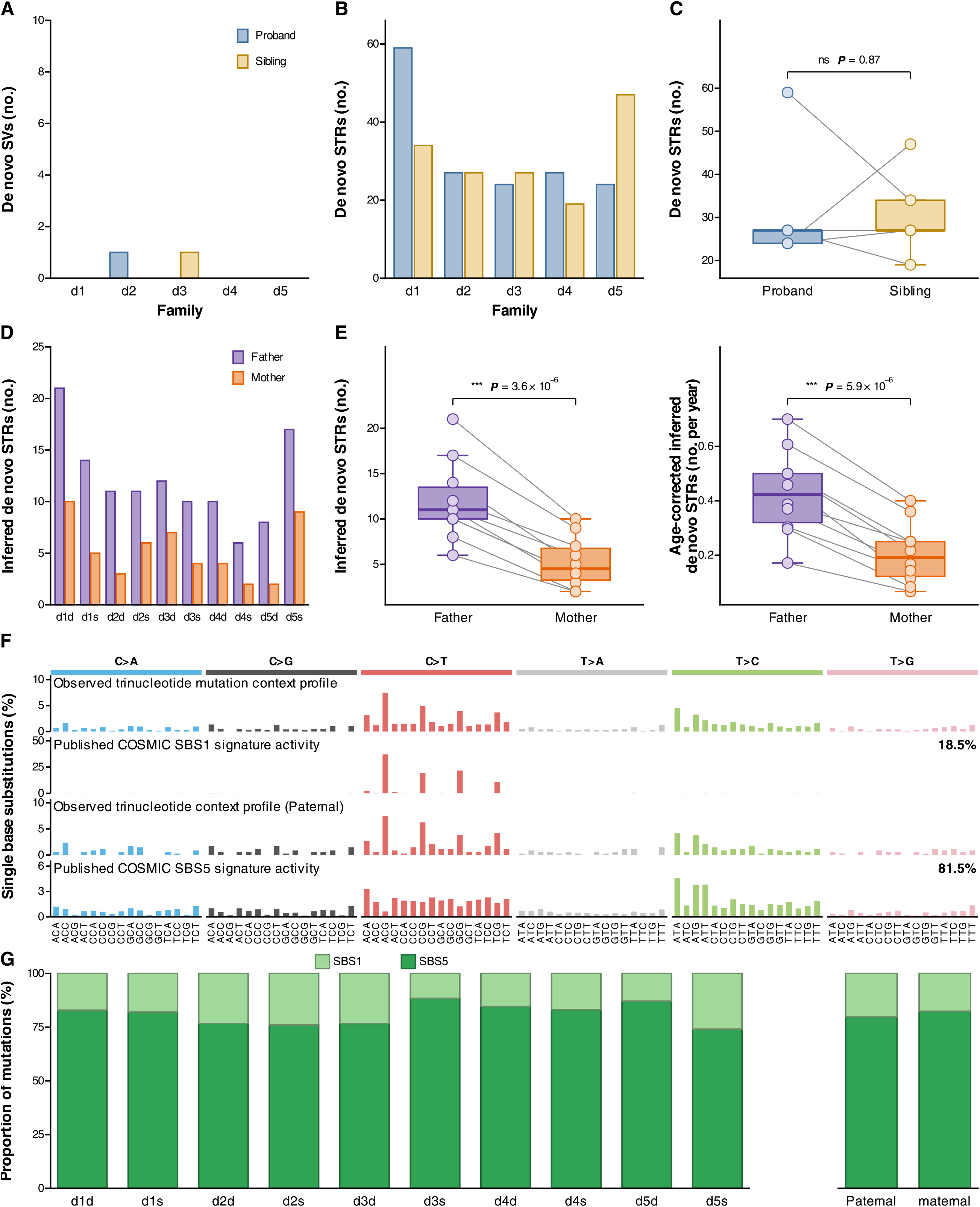
No excess in de novo Structural Variants (SVs), Short Tandem Repeats (STRs) or mutational signature processes in probands. de Novo SVs (A) and STRs (B) within the cohort. Only 2 de novo SVs are detected across the whole cohort. C) Comparison of de novo STR numbers between probands and siblings (Paired t-test, *P* = 0.866). D) Number of inferred parental origin of de novo STRs. E) Inferred (left) and age-corrected inferred (right) de novo STRs comparison between parents. (Paired t-test, *P* = 3.57E-06; *P* = 5. 87E-06). F) Trinucleotide mutation context profile from DNMs in child and paternal only transmitted DNMs (left). de novo mutations are attributed to SBS1 (clock-like spontaneous deaminase; ∼ number of divisions, 18.5%) and SBS5 (clock-like unknown; ∼ chronological time, 81.5%) cosmic signatures. G). Proportion de novo mutations from SBS1 and SBS5 cosmic signatures in probands and siblings (right) and from paternal and maternal origin (left). NS = not significant (P > 0.05). Three-star symbol (***) indicates P < 0.001.

Next, tandem repeat mutations were called using GangSTR^35^ and de novo mutations were inferred using a probabilistic method from MonSTR^36^. We detected 315 de novo STRs within the whole cohort (**Figure 4B**), with an average of 31.5 per child, but no excess in probands relative to their age-matched siblings (**Figure 4C**; *P*=0.87, paired t-test). Next, we assigned a parental origin to 172 out of the 315 de novo STRs (**Figure 4D**), showing a de novo STRs with preferential paternal transmission (**Figure 4E**) but no differences between probands and siblings.

Next, we tested whether the observed excess of de novo substitutions might be due to a mutational process with a distinctive trinucleotide signature. To address the low number of DNMs per child, we ran SigProfiler on the pooled mutations across probands and siblings for both mutational signature de novo extraction and for inference of the weights of the known COSMIC mutational signatures (**Methods**). This identified 18.5% of mutations as originating from COSMIC signature SBS1 (clock-like spontaneous deaminase; ∼ number of divisions) and 81.5% from SBS5 (clock-like unknown; ∼ chronological time)^37^ (**Figure 4F-G**). We could not attribute the excess to any other known or de-novo processes, suggesting that the excess might not be due to a distinct process in *TP53*-mutated probands or that the number of mutations might be too low to reliably identify a distinct etiology.

### A general model of post-zygotic newly acquired *TP53* mutations predicts unusually clonal *TP53* mutations in human cancers

A de novo mutation can arise at several stages: 1) pre-fertilisation, either as a constitutional or mosaic mutation in the parent, leading to a germline mutation in the proband; 2) post-fertilisation, as a post-zygotic mutation, leading to either constitutional or mosaic *TP53* in the proband (**Figure 5A-E**). A post-zygotic mutation reaches constitutional (near-heterozygous) allele fraction only if it arose in the zygote or first cleavage division^38^, or as we hypothesize, if it was subsequently swept to fixation by selection. To explain the observed low excess of de novo mutations, for which the spectra corresponds to known mutational processes (SBS1 and SBS5) on both parental copies, we emit three non-exhaustive hypotheses: 1) The excess causes the *TP53* mutation: an excess of DNM in the gametes of the parents would increase the chance of hitting *TP53*; 2) the presence of a *TP53* mutation causes the excess: a *TP53* mutation could lead to a slight excess of DNM in the parental gametes; 3) The excess is a byproduct of *TP53*-mutant clone selection in the child: an early post-zygotic *TP53* variants that swept through the population could lead to an excess of hitchhiking post-zygotic mutations.

**Figure 5.**
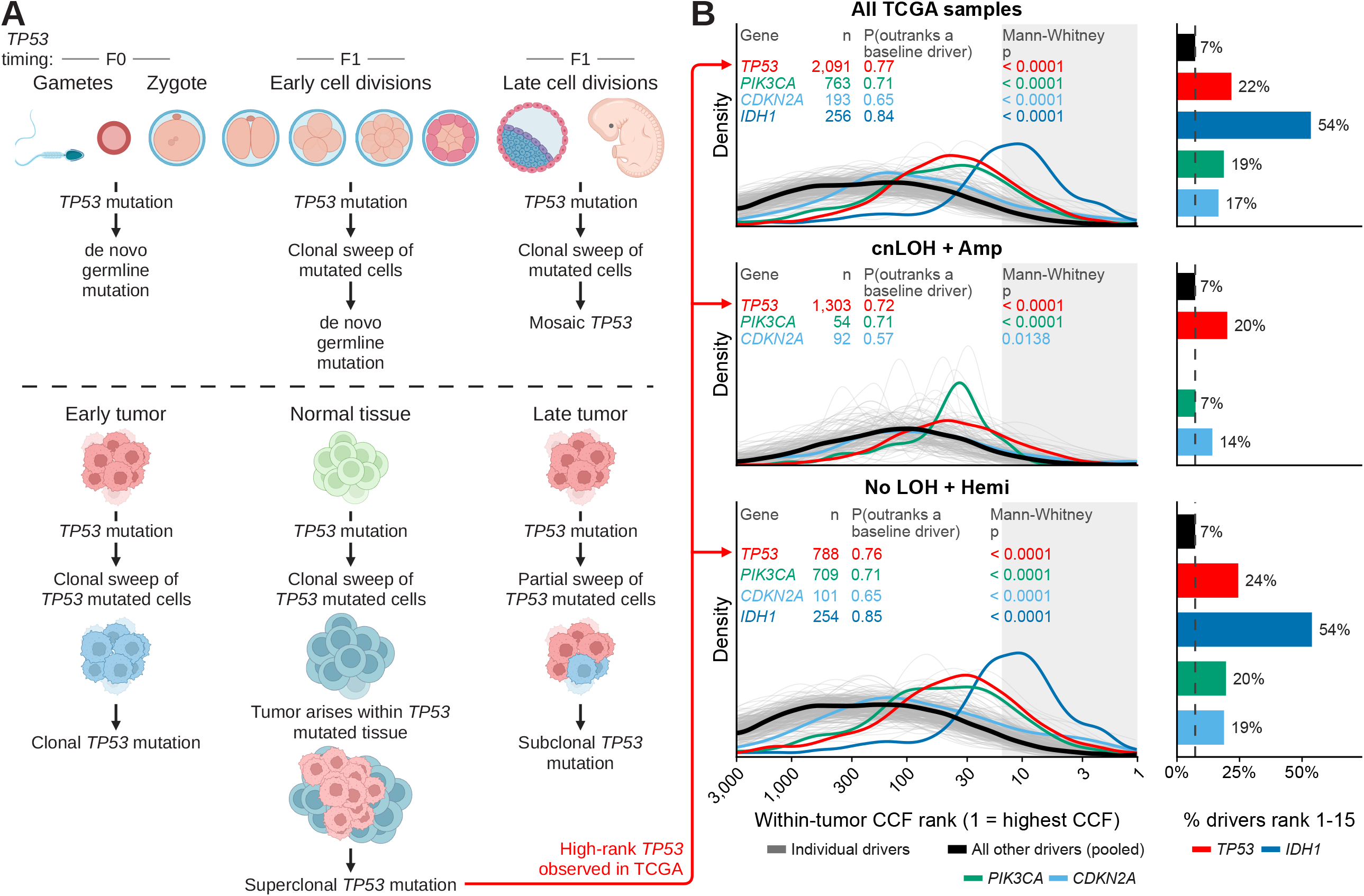
TP53 mutation timing leads to different heritability and cancer risk profiles. (A) Post-zygotic model of TP53 mutation acquisition in all cells. (B) Post-zygotic model of TP53 mutation acquisition in gonadal cells. (C) Post-zygotic model of TP53 mutation acquisition in somatic cells. (D) Inherited model of germline TP53 mutation. (E) Inherited model of TP53 mutation present only in the gametes of the parent. F) Schematic of the developmental and somatic contexts in which a *TP53* mutation can arise. Top (germline): a mutation occurring in a gamete or the zygote is transmitted as a true *de novo* germline mutation; a TP53 mutation arising during early cleavage (2-to 8-cell or morula stages) can drive a clonal sweep of the mutated lineage and likewise be observed as a *de novo* germline mutation; and a mutation arising later, at the blastocyst/early-embryo stage, populates only part of the soma and presents as mosaic *TP53*. Bottom (somatic): a mutation acquired at tumour initiation sweeps with the founding clone and is clonal; a mutation acquired in histologically normal tissue can sweep through the local cell population, so that the tumour arises within an already *TP53*-mutated field and the mutation is superclonal; and a mutation acquired after initiation occupies only a subset of cancer cells and is subclonal. The superclonal somatic scenario (bottom middle) is the somatic counterpart of the early-sweep germline scenario (top middle). (G) Within-tumour CCF rank of clonal driver mutations in 4,935 solid TCGA tumours. In each tumour, SNVs were ranked by descending CCF (rank 1 = highest, at right). Curves show rank densities of clonal mutations in *TP53* (red); *PIK3CA* and *CDKN2A*, the other drivers right-shifted in at least three tumour types (Supplementary Figure S6), and *IDH1*, the most right-shifted driver overall, which is right-shifted only in low-grade glioma; all other IntOGen driver genes pooled (black); and each of those genes separately (grey). Panels show all mutations (top) and mutations by copy-number state at the locus: copy-neutral or amplified LOH (middle) and no LOH or hemizygous loss (bottom). n, number of clonal mutations; P(outranks a baseline driver), probability that a mutation of the named gene ranks above a pooled driver mutation, from the Mann–Whitney U statistic (0.5 = no difference); p, two-sided Mann–Whitney test. Genes with fewer than 25 mutations in a panel are not shown. On the right, bars indicate the percentage of each gene’s mutations in ranks 1–15 (the shaded region on the density plot).

The first hypothesis suffers from reverse causation, as *TP53* status is used to define the groups, mutation burden conditional on that status cannot explain the origin of the *TP53* mutation. For the second hypothesis, in mice loss of Trp53 does not elevate spontaneous mutant frequencies in Trp53-null spermatocytes or spermatids relative to wild-type^39^. Finally, our third hypothesis, our results could be compatible with the *TP53* variants being preferentially post-zygotic (**Figure 5A-E**). Indeed, a *TP53* variant that happened in the early first divisions could lead to a sweep of the less *TP53*-competent lineage, as seen in *TP53*-/-mouse models of early embryogenesis^40,41^. The mutations that happened in the first cell divisions would hitchhike with the *TP53* variant and thus correspond to the observed excess.

We further posit that the timing of any post-zygotic TP53 variant can range from very early to very late. Later TP53 variants would have a lower allele frequency across cells, but be acquired at higher frequency in the population, i.e. across patients. A post-zygotic variant could thus lead to mosaic LFS for relatively early post-zygotic mutations to tiny clonal expansions restricted to single tissue compartments for later somatic or germline TP53 mutations. All of these could accrue the risk of developing a cancer within the TP53-mutated lineage. In support of our third hypothesis, we thus predict a similar phenomenon in human cancer development, where a significant fraction of the observed *TP53*-mutated cancers is initiated within larger *TP53*-mutated pre-cancer clones. *TP53*-mutant clones are common in normal adult epithelia: in middle-aged and older adults, *TP53* mutations are present in 5– 20% of oesophageal epithelial cells^42,43^, and an estimated ∼3% of oral epithelial cells carry a *TP53* driver mutation in adults aged 65–85^44^. If a tumour begins inside a *TP53-*mutated tissue, the initiating *TP53* mutation should be carried by every cancer cell and also by histologically normal cells adjacent to and admixed with the tumour. Because cancer cell fraction (CCF) is estimated from bulk DNA sequencing data based on the inferred cancer compartment, a mutation that is also present in the surrounding tissue would be present in more cells than the tumour compartment contains, resulting in a CCF that exceeds 1, hence called superclonal. If *TP53* clones are uniquely likely to sweep normal tissues later in life, *TP53* mutations in cancers should rank above the tumour’s other clonal mutations in estimated CCF (**Figure 5F**). Using the per-mutation CCF estimates from a recent high-accuracy reconstruction of thousands of whole exomes from TCGA^45,46^ and whole genomes from PCAWG^47^, we ranked somatic single-nucleotide variants within each tumour by descending CCF (rank 1 = most clonal) in 4,935 solid TCGA and 1,322 solid PCAWG tumours. As subclonal mutations should rank lower by definition, we restricted the comparison to mutations that CliPP assigned to each tumour’s clonal cluster, and asked how often a clonal *TP53* mutation outranked a clonal mutation drawn from a pooled baseline of other cancer driver genes (**Methods**). We assembled the comparator baseline from mutations in all other driver genes rather than from passenger mutations. Driver genes are likewise under positive selection and typically acquired early in tumour evolution, so excess *TP53* clonality relative to this baseline is less likely due to positive selection or early acquisition alone.

In TCGA, a clonal *TP53* mutation outranked a baseline driver mutation with probability 0.77 (Mann–Whitney P < 0.001), with one in five *TP53* mutations (21.8%) sitting within ranks 1– 15 for its tumour, compared with 7.3% of baseline driver mutations (**Figure 5G**). The result held when re-analyzing within copy-neutral or amplified LOH (0.72, Mann–Whitney P < 0.001) and within drivers without LOH or with hemizygous loss (0.76, Mann–Whitney P < 0.001). It also held within individual tumour types, where across TCGA *TP53* was right-shifted in 14 of 16 types (P = 0.59–0.71; **Supplementary Figure S6**). The only other drivers right-shifted in at least three tumour types, *PIK3CA* (0.71) and *CDKN2A* (0.65), ranked below *TP53*. In the pan-cancer view, *IDH1* ranked highest overall (0.84), but was right-shifted only in low-grade glioma (LGG). Notably, *NOTCH1* was also right-shifted within head and neck squamous cell carcinomas (P = 0.61, n = 34, within-type q = 0.047), with 18% of its mutations in ranks 1–15 compared with 9% of other driver mutations (**Supplementary Figure S6**), consistent with the frequent *NOTCH1*-mutant clones observed in normal oral epithelium^44^. In PCAWG, *TP53* again outranked the baseline in aggregate (0.57, Mann– Whitney P < 0.001) and in both copy-number states, while *PIK3CA* and *CDKN2A* did not (**Supplementary Figure S7**).

These observations are compatible with a model in which *TP53*-mutant clones expand before malignant transformation, such that some tumours arise within pre-existing *TP53*-mutant fields. More broadly, they support a continuum linking constitutional *TP53* mutations, mosaicism, normal-tissue clonal expansions, and cancer evolution through a common process of *TP53*-driven cellular selection.

## Discussion

The prevalence of LFS in the population is 1/3,555^48^ to 1/20,000^49^. Approximately 70% of LFS have a detectable constitutional pathogenic variant in *TP53*^50–52^. The remaining cases could be due to *de novo* germline, or mosaic *TP53* variants^53–56^, but current estimates likely remain lower bounds. Roughly one out of 6 LFS patients shows evidence of mosaic rather than constitutional *TP53*, akin to our m1 family^55,57^. Mosaic *TP53* mutations have important implications for both genetic counselling and patient management because they are post-zygotic. Indeed, compared to a pre-zygotic mutation, a post-zygotic origin fundamentally changes risk assessment for parents, relatives and for the offspring. Given the uncertain tissue distribution of mosaic mutations, affected individuals should be considered for *TP53*-adapted surveillance and, where feasible, avoidance of radiotherapy. Because the primordial germ cell lineage segregates early from other embryonic lineages, only sharing ∼8 SNVs with them^58^, the germ cells and therefore offspring of such mosaic cases would not be expected to harbour the variant. However, some mosaic variants can still be present in germline cells of the proband and remain transmissible to their offspring.

In theory, even a constitutional *TP53* mutation could be post-zygotic. While there is no evidence for positive selection of *TP53* mutations in either the paternal or maternal germ cells^59,60^, there is evidence for positive selection for somatic *TP53* variants in most tissues^40,44,61,62^; and for clonal sweeps of early *TP53*-/-mutants in embryogenesis in mice^40,41^. The timing of the post-zygotic *TP53* mutations determines whether it can sweep and become constitutional or instead lead to mosaicism. Some *TP53* mutations happening as early as during gastrulation still lead to mosaic rather than constitutional *TP53*^53^, thus a constitutional post-zygotic TP53 mutation would need to have appeared in the very early cell divisions.

So far, the origin of de novo pathogenic mutations in *TP53* has been unexplored. The rarity of de novo *TP53* mutations, combined with the stringent selection criteria applied here, resulted in a unique cohort. The availability of complete family quartets, including both parents, an affected proband, and an age-matched sibling, enabled a powerful controlled within-family study design. This framework substantially reduces confounding and provides an opportunity to investigate the developmental origins and genomic consequences of de novo *TP53* mutations. Thanks to both short- and linked-read sequencing of each genome, we found a significant excess of ∼15 DNM in the genomes of the probands despite the cohort of only 5 quartets. This excess did not seem to come from the paternal or maternal genomes exclusively, but the cohort size was too small to reliably assess parental-specificity of the excess.

To compare the pre- and post-fertilisation hypotheses, we reason that the number of mutations potentially hitting *TP53* would be roughly similar in the gametes (∼50-60 paternal, ∼5-20 maternal^25,31^) or in the first N cell divisions (2^(N-1) x mutation rate). For example, in the first 6 cell divisions assuming ∼2.5 mutations per division we would expect ∼15 mutations per cell and a total of ∼80 unique mutations across cells. In general, the probability of hitting *TP53* would be roughly proportional to ∼2^N, leading to exponentially higher chances of seeing smaller lineages (arising when N is large) within individuals, and non-zero chance of seeing clonal sweeps in early cell divisions (when N is very small) at the population level.

We propose that the deleterious de novo *TP53* variants in our Li-Fraumeni patients could be preferentially post-zygotic. Our data would then support a similar phenomenon in humans as seen in mice embryos^40^, where similarly to mice *TP53*-/-cells^41^, human *TP53*+/-cells could sweep during early cell divisions, such that the *TP53* mutations are shared with primordial germ cells and therefore heritable.

In many but not all normal human tissues, including epidermis, esophagus, blood or urothelium, or peritoneal tissues in women^63^, somatic evolution studies have shown *TP53* is a driver of local clonal expansions. *TP53* is also one of the most common pan-cancer drivers^64–67^ and in recent molecular timing analyses of cancer genomes, *TP53* is typically seen as an early clonal event with timing ranging from very early to close to biopsy^68^. Thus *TP53* can lead to sweeps both within early lineages and local tissue compartments. In contrast, *TP53* variants are not selected in sperm^59,60^ and make the sperm less motile^69,70^.

In theory a *TP53* pathogenic mutation can occur at any point in time or cell division, with an impact depending on its timing that would range from a full clonal sweep to a local clonal expansion. While the excess of DNM in our cohort might suggest early sweeps, we also found that *TP53* mutations in both TCGA^45,47^ and PCAWG^47^ were among the most clonal driver mutations, ranking above other clonal driver mutations in within-tumour CCF (**Figure 5G**; **Supplementary Figures S6 and S7**). A CCF greater than 1 suggests mutation acquisition in normal cells, in addition to tumour cells. While alternative explanations remain possible, these observations are compatible with somatic *TP53* variants acquired later with a localized sweep prior to tumour emergence, that is, with cancers arising within pre-existing *TP53*-mutant fields, and are consistent with reports that *TP53*-mutant clones are common in normal epithelium^42–44^.

Therefore, we propose an encompassing model where, depending on the timing of the acquisition of the *TP53* variant, the resulting clinical presentation can range from classical Li-Fraumeni syndrome with a constitutional *TP53* variant, and mosaic LFS to more local *TP53*-mutated clonal expansions often seen in cancer patients.

Our interpretation that a substantial fraction of the de novo *TP53* mutations are post-zygotic in de novo LFS patients remains speculative at this stage. However, it predicts that more cancer patients are in fact mosaic for *TP53*, ranging from cross-tissue mosaicism to within-compartment clonal expansions, and cancer patients with LFS are the extremes of a larger spectrum of *TP53*-mosaic patients. *TP53* with low allele frequencies are typically removed from germline calls, as they could represent clonal haematopoiesis rather than post-zygotic mutations^71,72^, therefore new approaches might be needed to revisit those calls at lower frequencies^73^.

Here, our unique quartet design with age-matched proband and sibling allowed us to control for parental age and differences in within-family mutation rates, allowing us to detect the excess. In the future, data on larger de novo *TP53* cohorts will help validate the excess we have observed; our model also predicts that re-analyses of *TP53* mutations at population scale should show high rates of “occult” *TP53* post-zygotic mosaicism.

## Methods

### Sequencing and data processing

Following collection of blood samples, DNA was extracted according to the manufacturer’s protocol. Whole-genome sequencing (WGS) was performed using 500ng DNA for each sample. DNA sequencing was performed with the 150bp length paired-end mode with Illumina HiSeq X sequencer, (mean depth = 35.79X, **Supplementary Figure S8**). In addition to WGS, a linked-read sequencing (LRS) was performed using 10X Genomics Chromium Genome technology and an Illumina NovaSeq 6000 sequencer.

Read quality of FASTQ files was assessed with FastQC (v0.11.9). Reads were aligned to the human reference genome V38 (hg38) using BWA-mem (v0.7.17) with default parameters. The reference file was downloaded from the Human genome reference builds from GATK. Downstream processing of BAM files was performed according to the Genome Analysis Toolkit (GATK) best practices (v4.2.3.0). Sequence alignment files were converted to sorted BAM files with SortSam from Picard (v2.26.4). PCR duplicates were detected and marked by MarkDuplicates (Picard, v2.26.4). Base quality score recalibration was performed using GATK v4.2.3.0.

### De novo mutation (DNM) calling

#### Single nucleotide variants & Small Insertions/Deletions

De novo single nucleotide variants (SNVs) and insertions/deletions (INDELs) were detected using four germline variant callers: HaplotypeCaller (GATK v4.2.3.0), DeepTrio (DeepVariant v1.8.0), DeNovoGear (v1.1.1) and Octopus (v0.7.4). Variant call sets initially generated with HaplotypeCaller and DeepTrio were combined and jointly genotyped using GenotypeGVCFs (GATK v4.2.3.0) and GLnexus (V1.4.1), respectively. A recalibration model was next built to score HaplotypeCaller variant quality using VariantRecalibrator (GATK v4.2.3.0). HaplotypeCaller variants were then filtered using ApplyVQSR following the recommended GATK pipeline. DeepTrio, DeNovoGear and Octopus were used with default parameters. De novo variants were subsequently identified according to the heterozygous genotype in child (0/1) and homozygous reference genotype in both parents (0/0). Variants with a variant allele frequency (VAF) lower than 0.27 or higher than 0.8 were filtered out for downstream analysis. The lower boundary was assessed by a one-sided binomial test against p = 0.5 (α = 0.01), which corresponds to a VAF threshold of 0.27 at the median depth of 15x.

A variant consensus was defined as a variant identified by at least three of the four mutation callers. Variants shared by two callers were manually reviewed using the Integrative Genome Viewer, IGV (v2.19.7). Consensus SNV variants were subsequently genotyped using alleleCount (v.4.3.0) by comparing the number of alternate alleles at each locus between tumour and matched normal samples.

#### Structural variants

Germline structural variant (SV) calling was performed with Delly (v1.1.6), Manta (v1.6.0), Smoove (v0.2.8), SvABA (v1.1.0) and GRIDSS (v2.2.13). Following SV calling, a first-pass filtering was applied on each sample using the respective SV caller threshold. de novo SV were identified according to their respective genotypes in children (0/1) and parents (0/0). Next, SURVIVOR (Jeffares 2017) was used to generate a consensus between each caller. Consensus variants were defined as variants identified by at least four out of the five SV callers. Following the consensus step, SVs were genotyped using the read depth-based tool SVTyper (v0.7.1). Since short reads sequencing leads to a high rate of false positive variants, SV variants were further filtered out and retained based on the following criteria : (1) a VAF between 0.27 and 0.8, (2) genotype quality > 20, (3) > 5 reads supporting the alternative allele; (4) > 6 reads supporting the reference allele, (5) a squared quality score > 200.

#### Short Tandem Repeats

Short tandem repeats (STRs) of 2 to 6 base pairs were detected with Tandem repeat callers GangSTR (v2.5.0GangSTR was ran with defaults parameters. GangSTR caller was provided with the tandem repeat catalog from Weisburg et al. (2026)^74^, containing 2,8 million tandem repeat loci. A locus-level filtering was applied using DumpSTR (TRTools, v6.1.0). Specifically, STRs were filtered with the following parameters, --gangstr-min-call-DP 20, --gangstr-max-call-DP 1000, --gangstr-filter-spanbound-only, --gangstr-filter-badCI, --min-locus-hwep 0.00001, --min-locus-callrate 0.8.

De novo STRs were identified using MonSTR (v1.0.0) with the following parameters --max-num-alleles 100, --output-all-loci, --min-num-encl-child 3 --max-perc-encl-parent 0.05, --min-encl-match 0.9, --min-total-encl 10, --posterior-threshold 0.5 --default-prior -3.

Briefly, MonSTR estimates the posterior probability of a de novo mutation occurring at each Tandem Repeat (TR) in each child using the following parameters: the genotype likelihoods of the parents and the child generated from GangSTR Tandem Repeat (TR) caller; the transmission probabilities for the four inheritance scenarios (0.25 each); a mutation transition probability describing an allele mutating by N repeat units; and a prior probability of mutation (10^-3^ mutations per locus per generation). Due to the high false positive rate in de novo STR detection, we applied additional filters. Variants with a posterior probability > 0.5 were retained, and homozygous de novo STR calls were excluded. Finally, we applied a stringent filtering strategy described in Goldberg et al. (2024)^75^. A schematic overview of the de novo mutation calling pipeline is shown in **Supplementary Figure S9**.

### Germline mutation calling

Germline mutations SNVs were detected using 3 germline variant callers: HaplotypeCaller (GATK v4.2.3.0), DeepVariant (v1.8.0) and Strelka2 (v2.9.10) while germline INDELs were called using HaplotypeCaller and DeepVariant only. HaplotypeCaller variants were filtered out as described in the DNM calling pipeline. For each caller, variants were selected if they met the following criteria: a minimum depth of coverage > 9, a variant allele frequency (VAF) > 0.349, and detected by all variant callers.

### Phasing

An inhouse pipeline was written in R (v4.3) to assess the parental origin of de novo variants using WGS and LRS data from each family. For each DNM, reads covering the DNM loci were extracted using SAMtools mpileup (v1.14) from the child alignment file. These reads were screened for single-nucleotide polymorphism (SNPs) with a heterozygous genotype in the child and that could be assigned to either the maternal or paternal haplotype based on parental genotype. The SNP database of parents was generated in each parental genome using HaplotypeCaller.

### Mutational signatures

The mutational process signature was detected using SigProfilerAssignment (V1.1.3) with the commands *sigProfilerExtractor* and *Analyze.cosmic_fit*.

#### TCGA and PCAWG superclonal analysis

Driver-mutation cancer cell fractions for 280 driver genes from 5,424 TCGA samples across 31 cancer types, and 314 driver genes from 2,008 PCAWG samples across 39 cancer types, were obtained from the download at https://bioinformatics.mdanderson.org/apps/CliPP/. From this resource we used the per-SNV CCF estimates and CliPP cluster assignments.

##### Cancer cell fraction

CCF was defined as the fraction of cancer cells in the sample carrying a given SNV, CCF = CP / ρ, where CP is cellular prevalence and ρ is sample purity, and was computed from the variant allele frequency f, purity ρ, local total copy number N_T_ and the inferred mutation multiplicity m as CCF = f / (m ρ) × (ρ N_T_ + 2(1 − ρ)).

##### Within-tumour CCF rank and driver-gene baseline

Within each tumour, somatic SNVs were ranked in descending order of CCF, so that rank 1 denotes the most clonal mutation in that tumour; ties were broken at random under a fixed seed. For TCGA the rank universe was all SNVs captured by the exome (median 101 per tumour). For PCAWG it was each tumour’s coding SNVs (median 43 per tumour), giving a rank scale on the same order as the exome. By comparing within-tumour ranks, rather than raw CCF values, each tumour serves as its own internal control, making the comparison more robust to sample level errors in purity and ploidy estimation, which rescale all mutations within a sample together and could otherwise produce an apparent excess of superclonal mutations.

Hematological tumour types were excluded (TCGA: LAML, DLBC; PCAWG: MALY, CLLE, CMDI, LAML, DLBC), as were mutations without a copy-number state. This left 4,935 TCGA tumours (29 tumour types) and 1,322 PCAWG tumours (34 types) with at least one clonal driver-gene SNV. Driver genes were taken from the IntOGen Compendium^76^. Four genes were examined individually: *TP53*; *PIK3CA* and *CDKN2A*, the only other drivers right-shifted in at least three tumour types in the within-tumour-type analysis below; and *IDH1*, the most right-shifted driver in the pan-cancer comparison. For each of these genes, the statistic was the probability that one of its mutations outranks a mutation drawn from the pooled remaining driver genes, with ties split (0.5 = no difference). This is the Mann–Whitney U statistic divided by the product of the two group sizes^77,78^, and p-values are from two-sided Mann–Whitney tests. Analyses were repeated by copy-number state at the locus: copy-neutral or amplified LOH (minor copy number 0, major ≥ 2) versus all other loci (no LOH, or hemizygous loss). Genes with fewer than 25 mutations in a stratum were not tested. Shaded windows in the figures (ranks 1–15 in TCGA, 1–4 in PCAWG) hold similar shares of baseline driver mutations (about 7%).

The same comparison was made within each TCGA tumour type with at least 25 *TP53* mutations and 200 driver mutations for the baseline, testing every driver gene with at least 30 mutations in that type against all other driver mutations in the type (190 gene × tumour-type tests), with Benjamini–Hochberg correction^79^ within type. A gene was called right-shifted in a tumour type when P > 0.5 and q < 0.05. To remove any outsized effect from hypermutators, colorectal, endometrial and gastric tumours were first split into POLE-mutant, MSI and microsatellite-stable groups using TCGA molecular subtypes, and any extreme outliers detected in the remaining cancer types were also excluded (n = 11).

### Statistical analysis

All computations were performed using R programming language (v4.3.0) and RStudio (2023.12.1.402). Figures were generated with ggplot2(v3.5.1)

## Data Availability

All data produced in the present study are available upon reasonable request to the authors

## Data and code availability

Raw sequencing data have been deposited in the dbGAP repository. The code used for data preprocessing, the pipeline, statistical analyses, and figure generation is available on GitHub (https://github.com/IRIBHM-computational-groups/LFS_DNM_excess_pipelines). Analyses were performed using R (v4.3).

## Acknowledgements

We also thank R. Lattuca for critical reading of the manuscript and code. MP was supported by Fund Doctor J.P. Naets managed by the King Baudouin Foundation. The results shown here are in part based upon data generated by the TCGA Research Network: https://www.cancer.gov/tcga.

## Author Contributions

M. Parmentier performed bioinformatics analyses and wrote the manuscript. M. Montierth performed the TCGA and PCAWG clonality analyses and wrote the manuscript. R. Li curated the LFS patient cohort and analyzed data, C. Vera Recio performed bioinformatics and statistical data analyses. P Chau performed Sanger sequencing experiments. B. Li advised on bioinformatics analyses. R. Chen supervised the linked-read sequencing. G. Lozano supervised the whole-genome sequencing data generation as well as Sanger sequencing experiments. M.Tarabichi and W. Wang supervised the project and wrote the manuscript. All authors have read and approved the manuscript.

---

**Supplementary Figure S1**: **Identification of TP53 and PALB2 mutations in m1 family proband.** A) The proband from m1 family harbours a TP53 mutation with a variant allele frequency (VAF) of 0.2 (left) which suggests a subclonal TP53 variant in the blood lineage (10 out 49 reads; *P* = 3.8E-05). Additionally, the patient also harbours a germline heterozygous variant in PALB2 gene (right). This variant is classified as pathogenic and is associated with an increasing risk of cancer. B) Lollipop plot depicting TP53 mutation location in TP53 gene and protein.

**Supplementary Figure S2:** (A) Identification of TP53 germline mutation in f1 and f2 families. (B) TP53 germline mutation location on TP53 gene and protein.

**Supplementary Figure S3:** (A) Total (left) and unique (right) number of germline SNVs and INDELs across the whole cohort (d1-d5 families). (B) Germline SNVs and INDELs per individual in d1-d5 families. (c) Same for f1 and f2 families. NS = not significant (P > 0.05)

**Supplementary Figure S4:** A) Number of de novo mutations for SNVs and INDELs in all chromosomes and autosomes only. Comparison of phaseable DNM count in autosomes between probands and siblings (Total: *P* = 0.317, MD = 5.20; Maternal: *P* = 0.665, MD = 0.80; Paternal: *P* = 0.249, MD = 4.40, paired t-test). NS = not significant (P > 0.05).

**Supplementary Figure S5:** (A) De novo mutations annotated for their pathogenicity according to The American College of Medical Genetics and Genomics (ACMG) classification in probands and siblings from d1-d5 families (left) and f1-f2 families (right). B) Same as (A) but per individual.

**Supplementary Figure S6: Within-tumour CCF rank of clonal driver mutations by TCGA tumour type.** Each tile shows one solid tumour type, labelled with its TCGA study abbreviation; colorectal (CRC), endometrial (UCEC) and gastric (STAD) tumours are restricted to their microsatellite-stable (MSS) subgroup. Within each tumour, all SNVs were ranked by descending cancer cell fraction (CCF), so rank 1 is the tumour’s highest-CCF mutation; the x-axis is log10-scaled with rank 1 at the right. Only mutations assigned by CliPP to the tumour’s clonal cluster are included. Curves are kernel densities of within-tumour ranks for *TP53* (red, shown in every tile), each other driver gene right-shifted in that tumour type (other colours), all remaining driver mutations in the type pooled (black) and each remaining driver gene with at least 10 mutations (grey). For each named gene, n is the number of clonal mutations and P is the probability that one of its mutations ranks above a mutation of another driver gene in the same tumour type (0.5 = no difference). q is from a two-sided Mann–Whitney test with Benjamini–Hochberg correction within the tumour type; an asterisk marks q < 0.05 after correction across all 190 gene × tumour-type tests. A gene is called right-shifted when P > 0.5 and q < 0.05. Shading marks ranks 1–15.

**Supplementary Figure S7: Within-tumour CCF rank of clonal driver mutations in PCAWG.** Data are from 1,322 solid PCAWG whole-genome tumours. Within each tumour, coding SNVs were ranked by descending cancer cell fraction (CCF), so rank 1 is the tumour’s highest-CCF coding mutation; the x-axis is log10-scaled with rank 1 at the right. Only mutations assigned by CliPP to the tumour’s clonal cluster are included. Panels show all mutations (top) and mutations by copy-number state at the locus: copy-neutral or amplified LOH (middle) and no LOH or hemizygous loss (bottom). Curves are kernel densities of within-tumour ranks for *TP53* (red), *PIK3CA* (green) and *CDKN2A* (light blue), all other IntOGen driver genes pooled (black) and each of those genes with at least 10 mutations separately (grey). For each named gene, n is the number of clonal mutations, P(outranks a baseline driver) is the probability that one of its mutations ranks above a pooled driver mutation (0.5 = no difference), and p is from a two-sided Mann–Whitney test. Genes with fewer than 25 mutations in a panel are not shown, including *IDH1* (21 clonal mutations) in every panel. On the right, bars indicate the percentage of each gene’s mutations in ranks 1–4 (approximately the same baseline percentile as ranks 1–15 in TCGA).

**Supplementary Figure S8: Sequencing quality metrics.** Whole genome sequencing: 35.7 +/-1.89X median coverage depth; 30 +/-0.45 median Phred score; 95.7+/-0.45 % median properly mapped reads. Linked-read sequencing: 7.35 +/-1.08X median coverage depth; 34.9+/-0.38 median Phred score; 82.95+/-2.93 % median properly mapped reads.

**Supplementary Figure S9: Mutation calling pipeline.**

