## Supplementary FigS1 for "Excess of de novo substitutions in the genomes of individuals with newly acquired germline *TP53* mutations"

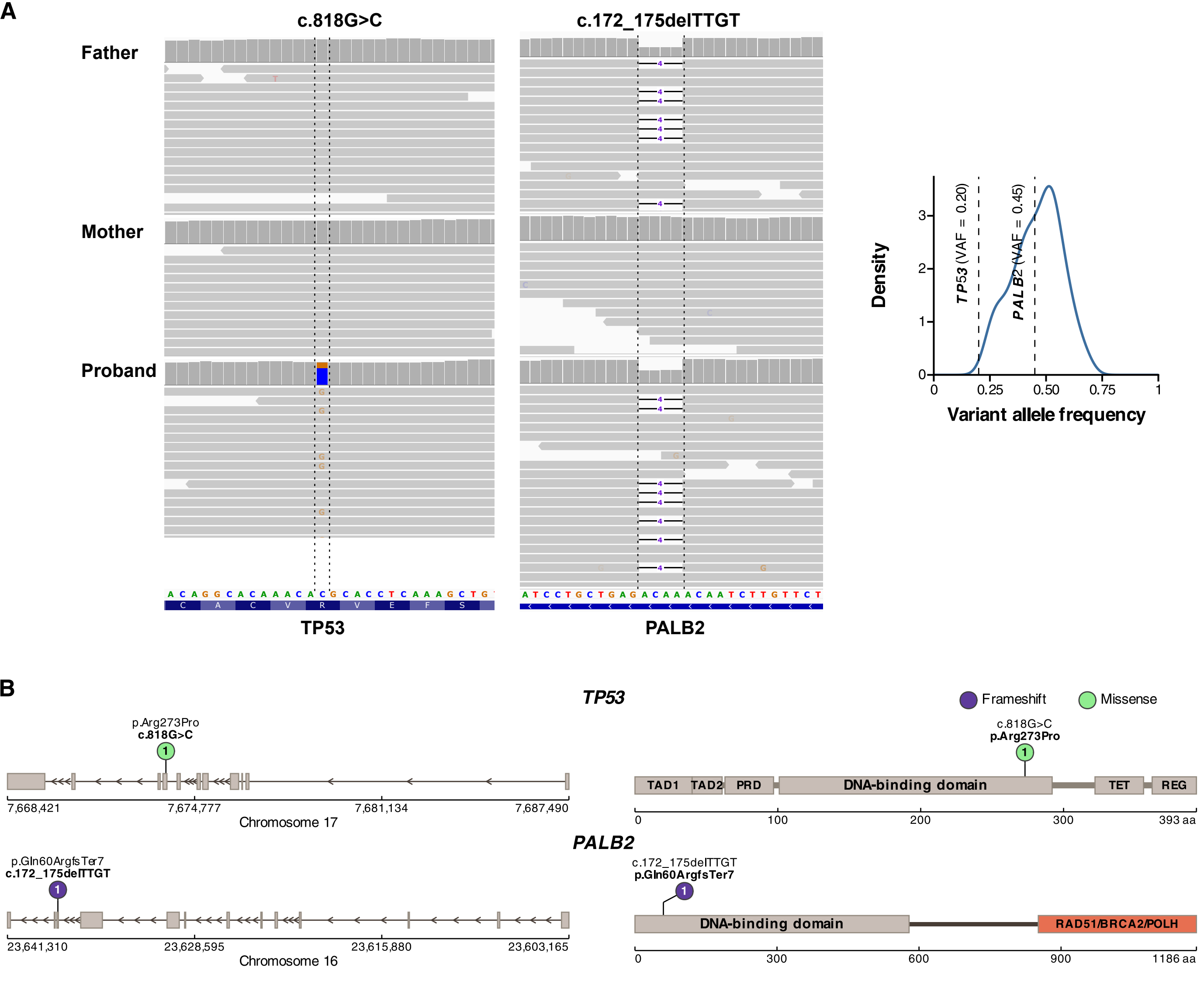

A
c.818G>C
c.172_175delTTGT
Father
Mother
Proband
TP53
PALB2
Density
TP53 (VAF = 0.20)
PALB2 (VAF = 0.45)
Variant allele frequency
0
0.25
0.50
0.75
1
B
TP53
Frameshift
Missense
p.Arg273Pro
c.818G>C
7,668,421
7,674,777
7,681,134
7,687,490
Chromosome 17
c.818G>C
p.Arg273Pro
TAD1
TAD2
PRD
DNA-binding domain
TET
REG
0
100
200
300
393 aa
PALB2
p.Gln60ArgfsTer7
c.172_175delTTGT
23,641,310
23,628,595
23,615,880
23,603,165
Chromosome 16
c.172_175delTTGT
p.Gln60ArgfsTer7
DNA-binding domain
RAD51/BRCA2/POLH
0
300
600
900
1186 aa
