## Supplementary FigS2 for "Excess of de novo substitutions in the genomes of individuals with newly acquired germline *TP53* mutations"

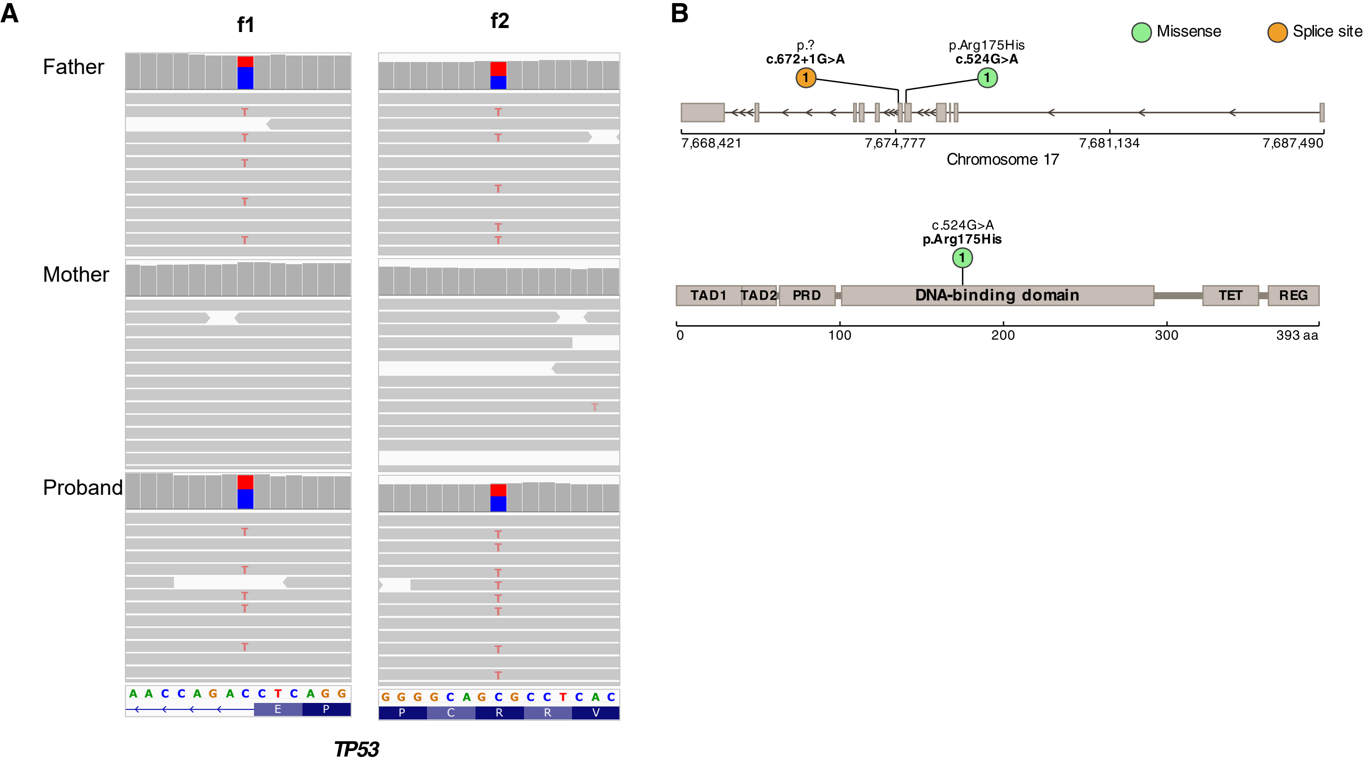
A
f1
f2
Father
Mother
Proband
A A C C A G A C C T C A G G
E P
G G G G C A G C G C C T C A C
P C R R V
TP53
B
Missense
Splice site
p.?
c.672+1G>A
p.Arg175His
c.524G>A
7,668,421
7,674,777
7,681,134
7,687,490
Chromosome 17
c.524G>A
p.Arg175His
TAD1
TAD2
PRD
DNA-binding domain
TET
REG
0
100
200
300
393 aa
