## Supplementary FigS3 for "Excess of de novo substitutions in the genomes of individuals with newly acquired germline *TP53* mutations"

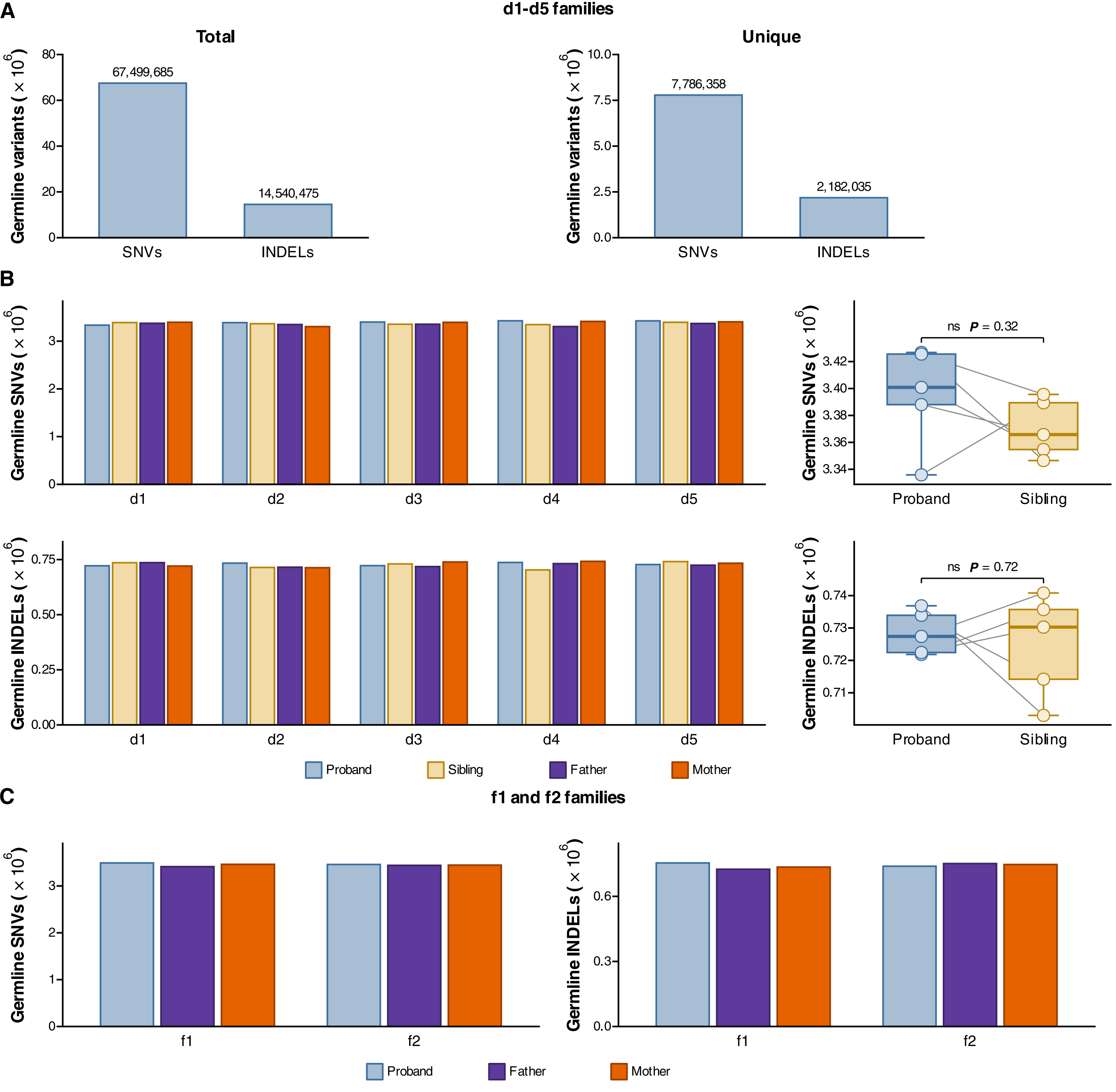

A
d1-d5 families
Total
Unique
Germline variants ( × 10^6)
67,499,685
14,540,475
7,786,358
2,182,035
SNVs
INDELs
B
Germline SNVs ( × 10^6)
Germline INDELs ( × 10^6)
d1
d2
d3
d4
d5
ns P = 0.32
ns P = 0.72
Proband
Sibling
Father
Mother
C
f1 and f2 families
f1
f2
