## Supplementary FigS4 for "Excess of de novo substitutions in the genomes of individuals with newly acquired germline *TP53* mutations"

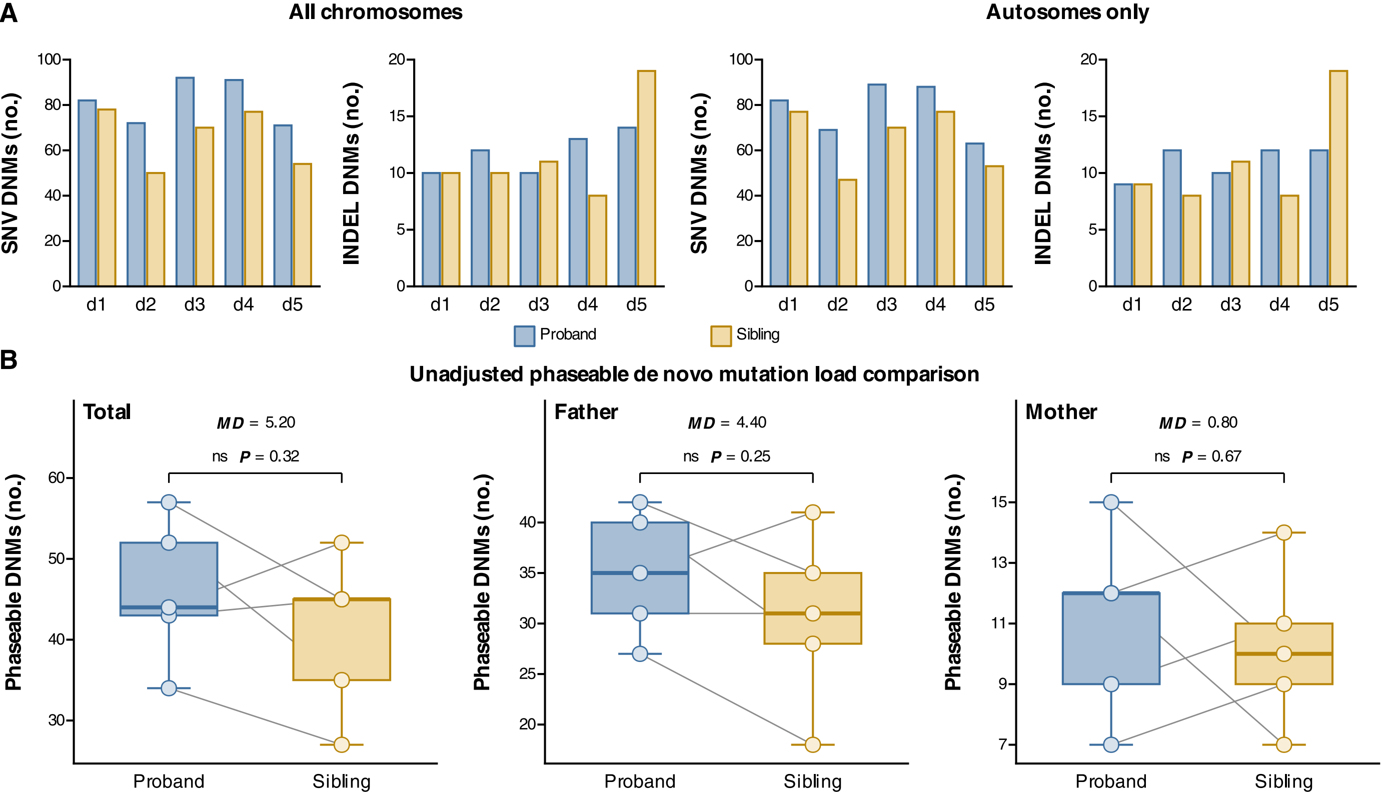
A
All chromosomes
Autosomes only
SNV DNMs (no.)
INDEL DNMs (no.)
d1
d2
d3
d4
d5
Proband
Sibling
B
Unadjusted phaseable de novo mutation load comparison
Total
MD = 5.20
ns P = 0.32
Father
MD = 4.40
ns P = 0.25
Mother
MD = 0.80
ns P = 0.67
Phaseable DNMs (no.)
Proband
Sibling
