## Supplementary FigS5 for "Excess of de novo substitutions in the genomes of individuals with newly acquired germline *TP53* mutations"

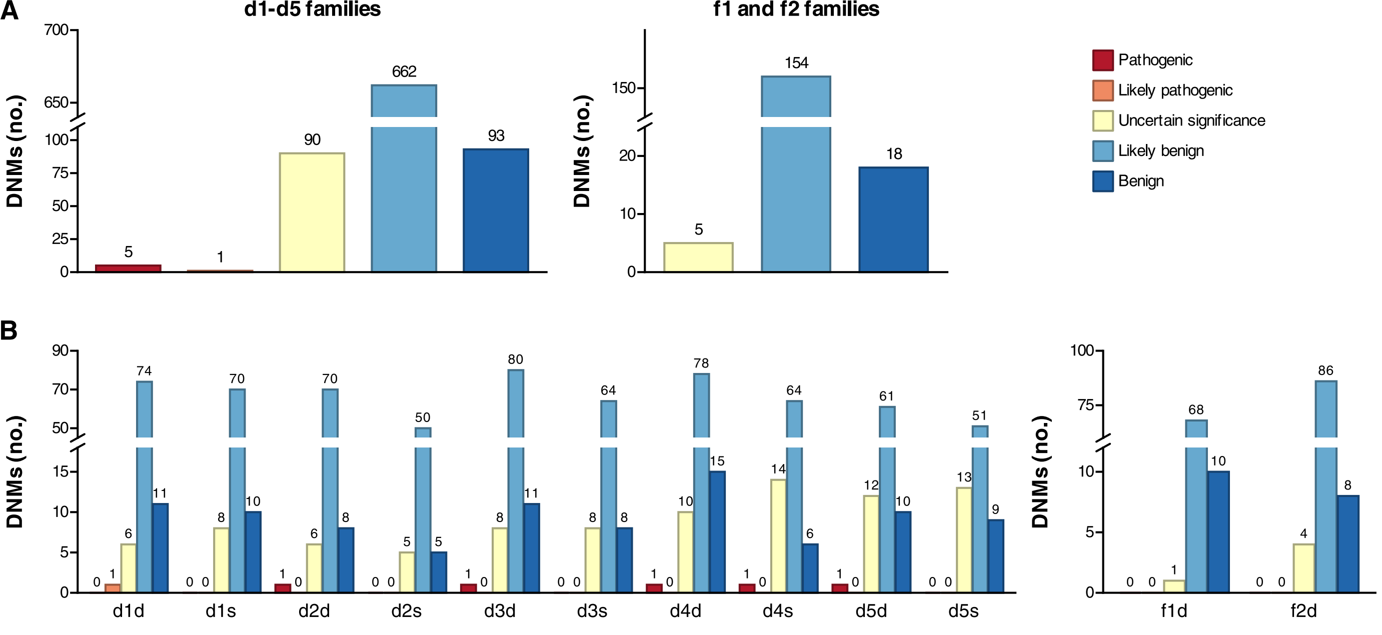
A
d1-d5 families
f1 and f2 families
DNMs (no.)
700
650
100
75
50
25
0
5
1
90
662
93
150
20
10
5
154
18
Pathogenic
Likely pathogenic
Uncertain significance
Likely benign
Benign
B
90
70
50
15
10
5
0
100
75
d1d 0 1 6 74 11
d1s 0 0 8 70 10
d2d 1 0 6 70 8
d2s 0 0 5 50 5
d3d 1 0 8 80 11
d3s 0 0 8 64 8
d4d 1 0 10 78 15
d4s 1 0 14 64 6
d5d 1 0 12 61 10
d5s 0 0 13 51 9
f1d 0 0 1 68 10
f2d 0 0 4 86 8
