## Supplementary figures and images for "Excess of de novo substitutions in the genomes of individuals with newly acquired germline *TP53* mutations"

### Supplementary FigS6

TP53 and every other right-shifted driver, within each solid TCGA tumor type

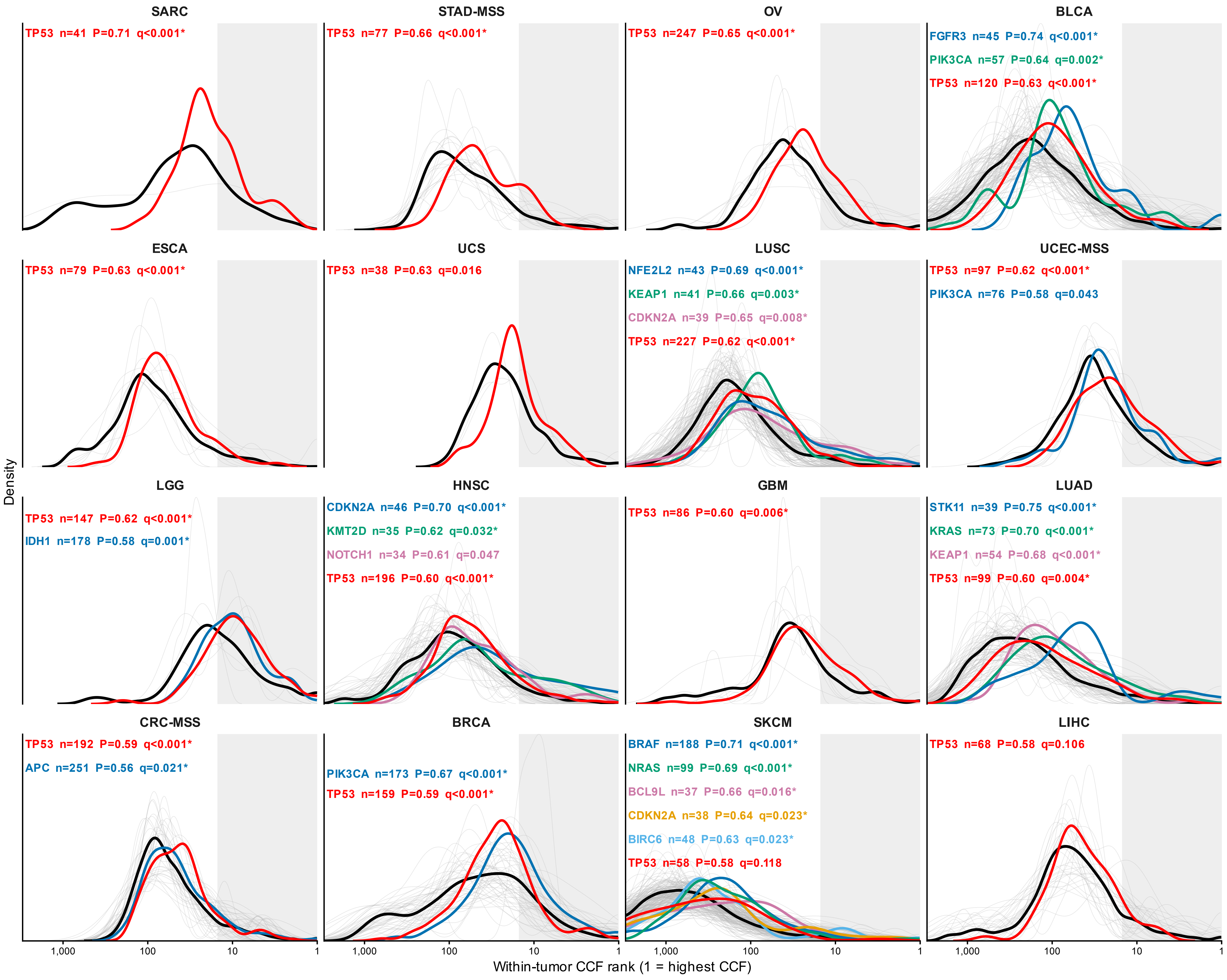
