## Supplementary FigS7 for "Excess of de novo substitutions in the genomes of individuals with newly acquired germline *TP53* mutations"

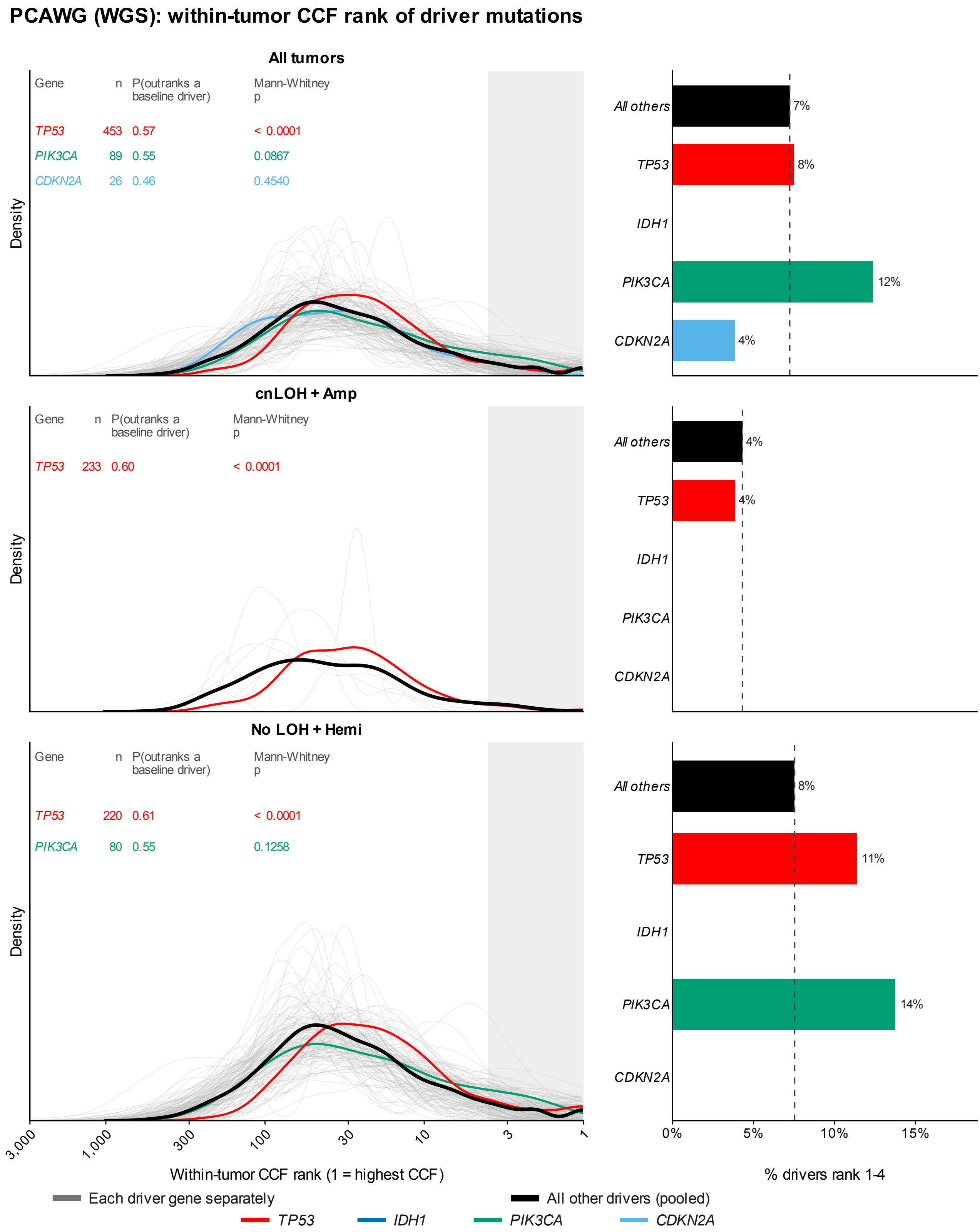

PCAWG (WGS): within-tumor CCF rank of driver mutations
All tumors
Gene
n
P(outranks a baseline driver)
Mann-Whitney p
TP53 453 0.57 < 0.0001
PIK3CA 89 0.55 0.0867
CDKN2A 26 0.46 0.4540
Density
All others
7%
TP53
8%
IDH1
PIK3CA
12%
CDKN2A
4%
cnLOH + Amp
Gene
n
P(outranks a baseline driver)
Mann-Whitney p
TP53 233 0.60 < 0.0001
Density
All others
4%
TP53
4%
IDH1
PIK3CA
CDKN2A
No LOH + Hemi
Gene
n
P(outranks a baseline driver)
Mann-Whitney p
TP53 220 0.61 < 0.0001
PIK3CA 80 0.55 0.1258
Density
All others
8%
TP53
11%
IDH1
PIK3CA
14%
CDKN2A
3,000
1,000
300
100
30
10
3
1
0%
5%
10%
15%
Within-tumor CCF rank (1 = highest CCF)
% drivers rank 1-4
Each driver gene separately
All other drivers (pooled)
TP53
IDH1
PIK3CA
CDKN2A
