## Supplementary FigS8 for "Excess of de novo substitutions in the genomes of individuals with newly acquired germline *TP53* mutations"

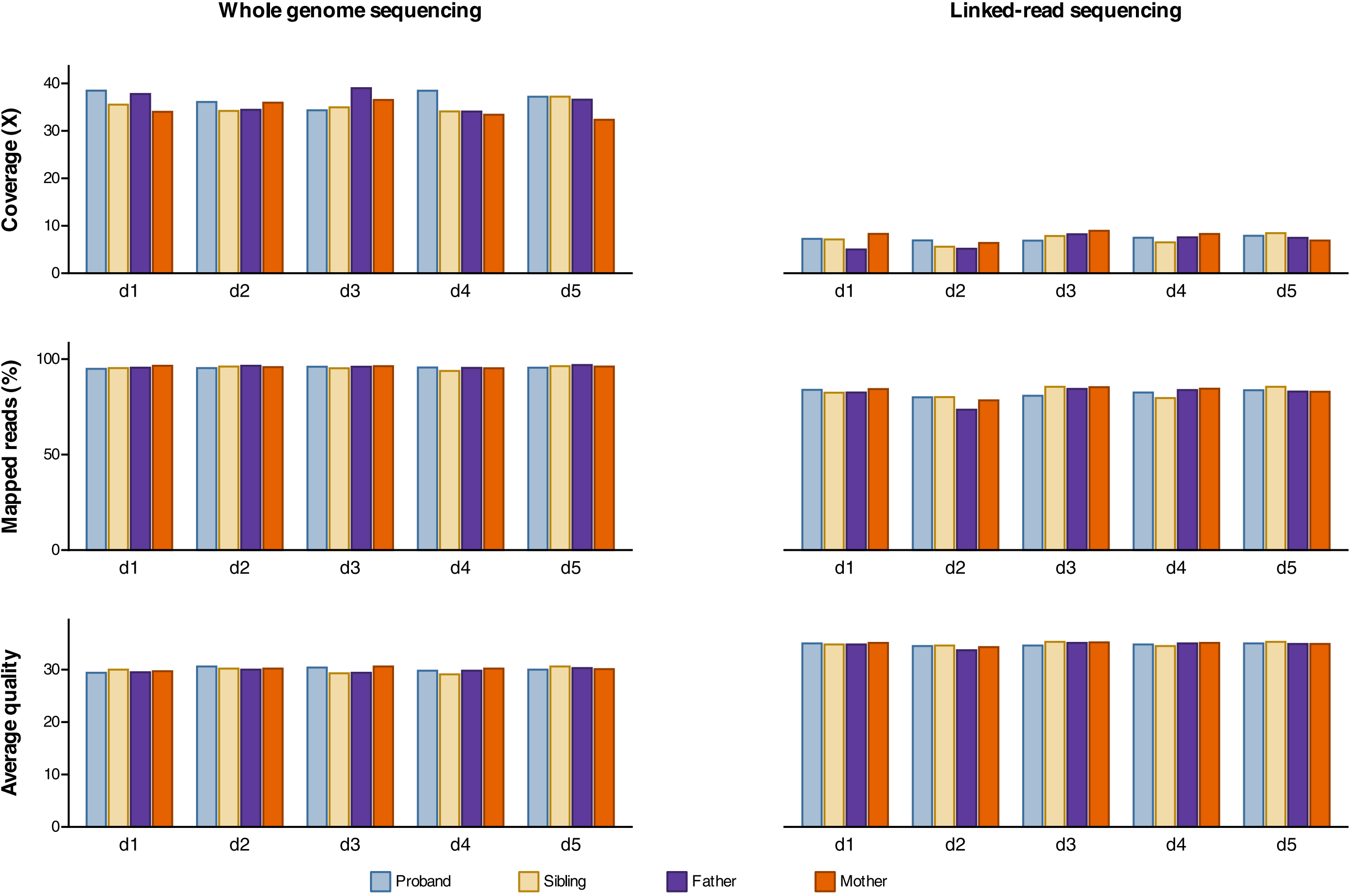

Whole genome sequencing
Linked-read sequencing
Coverage (X)
Mapped reads (%)
Average quality
0
10
20
30
40
50
100
d1
d2
d3
d4
d5
Proband
Sibling
Father
Mother
