## Supplementary FigS9 for "Excess of de novo substitutions in the genomes of individuals with newly acquired germline *TP53* mutations"

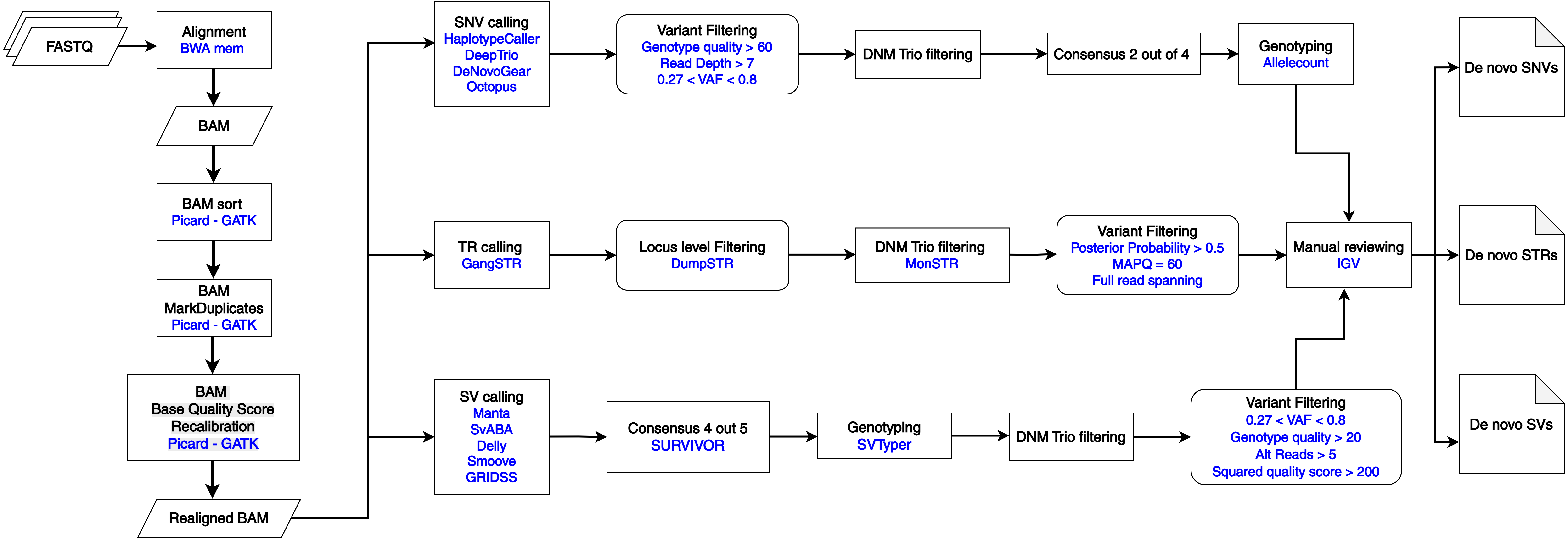
FASTQ
Alignment
BWA mem
BAM
BAM sort
Picard - GATK
BAM
MarkDuplicates
Picard - GATK
BAM
Base Quality Score
Recalibration
Picard - GATK
Realigned BAM
SNV calling
HaplotypeCaller
DeepTrio
DeNovoGear
Octopus
Variant Filtering
Genotype quality > 60
Read Depth > 7
0.27 < VAF < 0.8
DNM Trio filtering
Consensus 2 out of 4
Genotyping
Allelecount
TR calling
GangSTR
Locus level Filtering
DumpSTR
DNM Trio filtering
MonSTR
Variant Filtering
Posterior Probability > 0.5
MAPQ = 60
Full read spanning
Manual reviewing
IGV
SV calling
Manta
SvABA
Delly
Smoove
GRIDSS
Consensus 4 out 5
SURVIVOR
Genotyping
SVTyper
DNM Trio filtering
Variant Filtering
0.27 < VAF < 0.8
Genotype quality > 20
Alt Reads > 5
Squared quality score > 200
De novo SNVs
De novo STRs
De novo SVs
